# Fine-tuned Medical Vision–Language Model (MedGemma 1.5 4B) with and without Structured Radiologist Descriptors versus EU-TIRADS for Malignancy Risk Stratification of Thyroid Nodules at Ultrasound: A Tunisian Multicenter Diagnostic Accuracy Study

**DOI:** 10.64898/2026.10.21.26366156

**Authors:** Mohamed Nour Bou Ali, Najoua Lassoued, Houssem Mahmoud, Emna Mnif, Slim Haouet, Khaled Ben Rhouma, Mohamed Khaled Elsakly, Mehdi Hasnaoui

## Abstract

**Purpose:** To evaluate a medical vision–language foundation model fine-tuned on thyroid ultrasound images, with or without the radiologist’s structured descriptors, against expert-rated EU-TIRADS for the classification of thyroid nodules as benign or malignant.

**Materials and Methods:** This retrospective study included nodules that underwent ultrasound-guided fine-needle aspiration (FNA) between January 2019 and August 2025 in two Tunisian centers, with consecutive recruitment, together with a selection of images from an international public dataset. Of 1,580 nodules, 1,456 (272 malignant; 1,280 patients) with an interpretable cytological or histological reference standard were allocated at patient level to training (n = 942), validation (n = 258) and a test set sealed before training (n = 256; 51 malignant). MedGemma 1.5 4B was fine-tuned with low-rank adaptation under four input configurations: the image with nodule biometry, with or without age and sex (A, B); A with chain-of-thought supervision (C); and A plus the eleven qualitative descriptors of the radiologist’s structured report (D). The EU-TIRADS category was never a model input. Areas under the receiver operating characteristic curve (AUROCs) were compared with DeLong’s test and a prespecified non-inferiority margin of 0.05; EU-TIRADS ≥ 4 was the prespecified comparator, with ≥ 5 as a sensitivity analysis.

**Results:** Config D reached an AUROC of 0.953 (95% CI 0.909–0.976), versus 0.948 (0.894–0.972) for EU-TIRADS (difference +0.004, 95% CI −0.016 to +0.025), which met the non-inferiority criterion; Configs A–C, without the qualitative descriptors, reached 0.883–0.892 (Holm-adjusted p = 0.045 versus EU-TIRADS). At ≥ 4, EU-TIRADS had a sensitivity of 98.0% and produced 94 false positives among 205 benign nodules; at a threshold fixed on validation data, Config D reclassified 72 of them as negative while missing two additional cancers. Against EU-TIRADS ≥ 5 (sensitivity 92.2%, specificity 95.1%), Config D at its native threshold (88.2%, 94.1%) did not classify more nodules correctly (McNemar p = 0.22), and at its two other thresholds it classified fewer nodules correctly (p = 0.004 and 0.007). Raw probabilities were miscalibrated (expected calibration error 0.117 for Config D), and Platt scaling within the test set reduced this error to 0.019–0.040.

**Conclusion:** Given the ultrasound image together with the radiologist’s structured descriptors, the fine-tuned model matched expert EU-TIRADS discrimination and, against the ≥ 4 threshold, reclassified most false positives as negative, a gain concentrated in category 4 and based on only three cancers in category 4. It is best positioned as a report-standardizing second reader in the EU-TIRADS 4 gray zone, pending prospective external validation.

**Summary Statement:** A fine-tuned medical vision–language model given the ultrasound image and the radiologist’s structured report matched expert EU-TIRADS discrimination of thyroid nodules and would have deferred most aspirations of benign category 4 nodules, at the cost of one or two missed category 4 cancers depending on the threshold, based on only three cancers in category 4.

**Key Points:**

- On a test set of 256 thyroid nodules sealed before training, the model given the ultrasound image and the radiologist’s 16-field structured report reached an area under the receiver operating characteristic curve of 0.953, non-inferior to expert EU-TIRADS (0.948).
- Against EU-TIRADS 4 or higher, the model reclassified as negative 72 of the 94 false-positive nodules at a threshold fixed on validation data, at the cost of two additional missed cancers.
- Adding the radiologist’s qualitative descriptors to the image raised the area under the curve from 0.892 (Config A) to 0.953, and Platt scaling reduced the calibration error of the model from 0.117 to 0.019–0.040.

## Introduction

High-resolution cervical ultrasound detects thyroid nodules in a large proportion of adults [1], yet cancer is found in only 1–5% of nodules in unselected populations [2]. Risk stratification systems translate sonographic features into an ordered risk category that guides the decision to perform fine-needle aspiration (FNA). In Europe, EU-TIRADS is the reference framework [3] and is endorsed by the 2023 European Thyroid Association guidelines [2]. These systems share a characteristic profile: high sensitivity obtained at the price of limited specificity, so that many aspirated nodules turn out to be benign. Reported rates of unnecessary biopsy range from 25% to 55% depending on the system [4], and diagnostic odds ratios range from 2.2 to 4.9 across five systems [5]. Interobserver agreement is moderate for the risk category and only fair for individual descriptors such as echogenicity, shape and margins [6,7], and category 4 behaves as a gray zone for decision making.

Epidemiology adds weight to this imprecision. Thyroid cancer incidence has risen worldwide while mortality has remained flat, a pattern attributed largely to overdiagnosis driven by imaging [8,9]. Northern Tunisia follows the same course: in women, the age-standardized incidence recorded by the regional cancer registry rose from 3.7 per 100,000 in 1994 to 6.9 in 2010–2014 and is estimated at 9.0 in 2023 [10], and incident thyroid cancer cases in North Africa and the Middle East increased by 396% between 1990 and 2019 [11]. A tool able to remove benign nodules from the aspiration pathway without missing cancers would therefore address a growing problem.

Artificial intelligence (AI) has been applied to this task for more than a decade. Machine learning models trained on ultrasound images, whether radiomic or deep learning based, reach summary areas under the receiver operating characteristic curve (AUROCs) of about 0.92 in meta-analyses [12,13], and multicenter systems such as ThyNet [14] and the multimodal ThyGPT [15] reduced unnecessary FNA in large Chinese cohorts. Multimodal medical foundation models such as BiomedCLIP [16], LLaVA-Med [17] and MedGemma [18,19] now combine a domain-pretrained vision encoder with a language decoder and can be adapted to a specific task by parameter-efficient fine-tuning, for example low-rank adaptation (LoRA) [20,21]. Their appeal in settings with limited resources is practical: the weights are open, training fits on a single graphics processing unit, and no proprietary application programming interface is needed. Independent evaluations of general-purpose multimodal language models on thyroid ultrasound have, however, reported variable and often modest accuracy [22,23], which calls for task-specific evaluation.

Three gaps motivated this study. Most AI evidence in thyroid ultrasound comes from East Asian or North American cohorts, whose sonographic and histological case mix may not transfer to Maghrebi populations, and a recent meta-analysis restricted to studies with external datasets explicitly called for validation outside Asia [12]. Evaluations frequently report discrimination alone, without calibration or clinical utility, and often without a test set sealed before any training decision [24,25]. Finally, when an AI system is compared with a radiological score, the score itself is sometimes given to the model, which makes the comparison circular [26]. Supplying the descriptors from which the score is derived, or using the score as a training target, creates a partial form of the same problem, and its size has to be estimated.

We fine-tuned MedGemma 1.5 4B with LoRA to classify thyroid nodules as benign or malignant on B-mode ultrasound images in a Tunisian multicenter cohort, and compared it with EU-TIRADS, as rated by the radiologists of each source, on the same nodules of a test set locked by SHA-256 checksum before training. The EU-TIRADS category itself was never supplied to the model. We hypothesized that the fine-tuned model would reach an AUROC of at least 0.89 and would not be inferior to EU-TIRADS by more than 0.05.

## Materials and Methods

### Design, setting and reporting

This retrospective multicenter diagnostic accuracy study covered 1 January 2019 to 1 August 2025, with consecutive recruitment in the two Tunisian centers and a selection of images from an international public dataset. Reporting follows STARD 2015 [27,28], TRIPOD+AI [29], TRIPOD-LLM [30] and CLAIM [31,32]; the completed checklists are provided in Supplementary Tables S1–S3. The study adhered to the Declaration of Helsinki [33] and was approved by the Medical Ethics Committee of Taher Sfar University Hospital, Mahdia (reference CEM-2025-10-04). Because the data were retrospective and pseudonymized, individual consent was waived under Tunisian regulations (decree no. 2007-2609), and data processing complied with Tunisian organic law no. 2004-63 on personal data protection. No data left the country; all computation was performed locally on site in Tunisia, where the engineering co-author worked.

In STARD terms, the index test was the malignancy probability returned by the fine-tuned model, the reference standard was Bethesda cytology or, when available, definitive surgical histology, and the comparator was the EU-TIRADS category assigned in routine practice (for the public dataset, assigned by the study radiologists without knowledge of the pathology result).

### Participants and sources

Twenty-six radiology and pathology units across five Tunisian governorates were approached. Two centers met all eligibility criteria (a certified interventional ultrasound unit, a pathology laboratory issuing structured Bethesda reports, and joint participation of both services) and agreed to take part.

#### Source 1, Mahdia (Taher Sfar University Hospital catchment area)

Patients followed in endocrinology and referred for ultrasound-guided FNA; imaging and EU-TIRADS rating by a radiologist with at least 20 years of experience, and cytopathology by an affiliated laboratory.

#### Source 2, international public dataset

To widen the range of sonographic appearances seen in training, a selection of images was drawn from the open dataset of Hou et al. (8,508 images with binary histological annotation; CC BY 4.0) [34]. Images were selected for the sharpness of the nodule and the overall image quality, with a malignancy prevalence close to that of the Tunisian cohort as a target. The reference standard for these nodules is the histological diagnosis supplied with the dataset, and no cytological category applies. The images were re-interpreted locally: one of two radiologists with 5 years of experience, blinded to the pathology result, assigned the EU-TIRADS category, measured the nodule and recorded the eleven qualitative descriptors according to the 2017 EU-TIRADS standard [3]. Images were cropped and resized to the native resolution of the vision encoder, as the license permits.

#### Source 3, Tunis (CERU, Place Pasteur, and an affiliated pathology laboratory)

A national tertiary referral practice; imaging and EU-TIRADS rating by a radiologist with 30 years of experience. Because this center receives complex referrals from the whole country, a spectrum effect on sensitivity, specificity and predictive values was expected; it is addressed in the Discussion [35].

#### Eligibility

Inclusion required an age of 18 years or older at FNA; at least one nodule rated EU-TIRADS 2–5 that underwent ultrasound-guided FNA; an interpretable Bethesda report (categories II–VI) [36]; and a high-resolution image of the target nodule in at least one orthogonal plane showing at least 80% of the region of interest. Because aspiration was the entry criterion, nodules aspirated outside size-based guideline indications, including EU-TIRADS 2 nodules, were eligible. In patients with several nodules, only cytologically verified nodules were included. Exclusion criteria were nondiagnostic cytology (Bethesda I), previous thyroid surgery or cervical irradiation, previous thermal or ethanol ablation or radioiodine therapy, FNA not guided by ultrasound, and insufficient image quality. The documented nodules are those that met these initial eligibility criteria; nodules with nondiagnostic cytology were excluded a priori.

### Reference standard

A two-tier reference standard was used. In the Tunisian centers, Bethesda II defined a benign outcome and Bethesda V or VI a malignant outcome [36]; when surgical histology was available in the pathology records, it superseded cytology. For the public dataset, the reference standard was the histological diagnosis supplied with the images. Nodules with indeterminate cytology (Bethesda III–IV) were included only if postoperative histology was available; the other indeterminate nodules were excluded before the partition and contributed to neither training nor evaluation (Figure 1). Requiring histology for every nodule would have introduced a major verification bias, because only surgically treated, and therefore higher-risk, nodules would have been verified. The consequences of this choice were estimated with a Begg–Greenes reweighting (Supplementary Appendix) [37].

**Figure 1.**
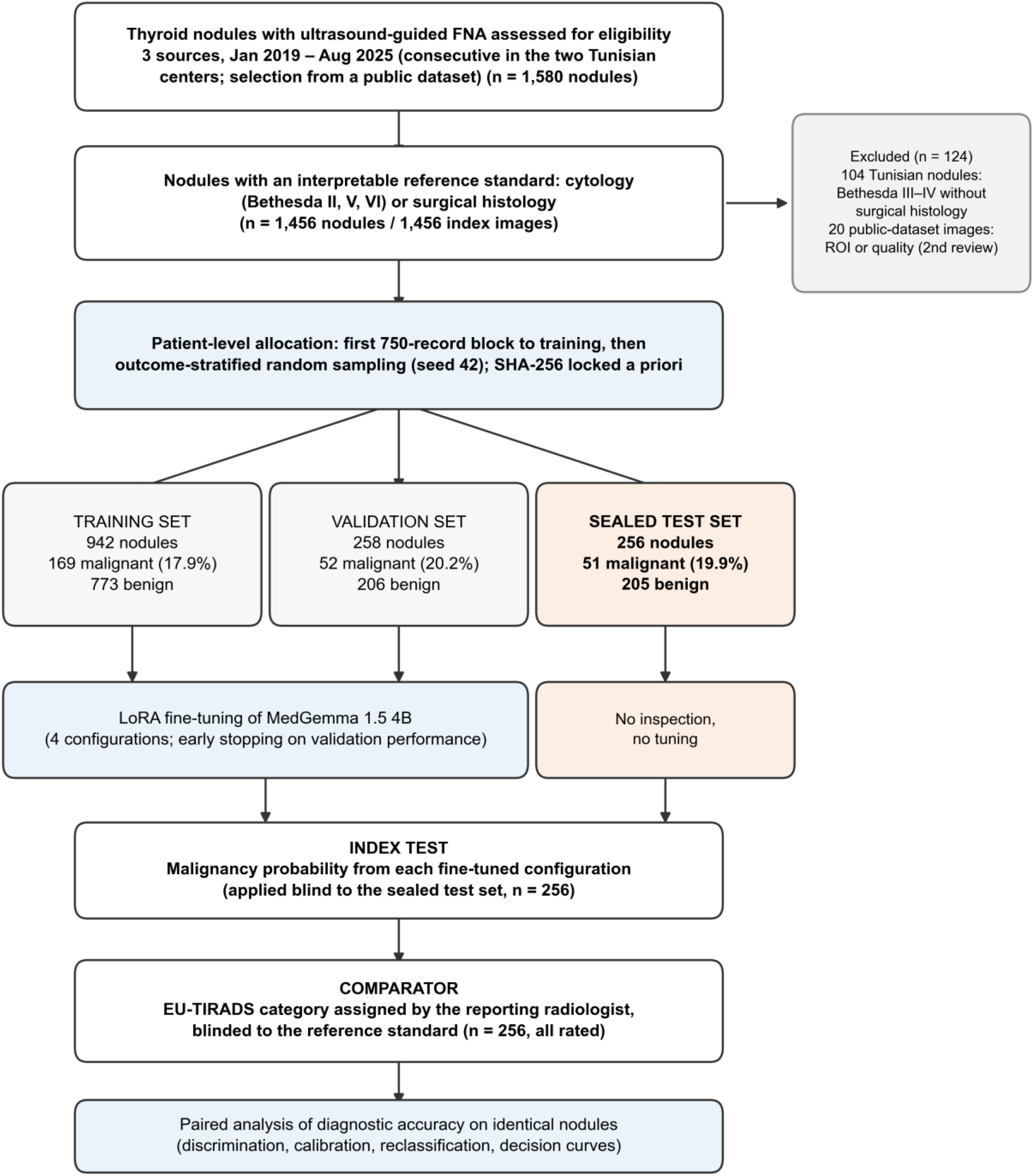
Study flow diagram (STARD 2015). Flow from the 1,580 documented nodules, which met the initial eligibility criteria (consecutive in the two Tunisian centers; nondiagnostic Bethesda I cytology excluded a priori), to the sealed test set. The 124 excluded nodules (104 Tunisian nodules with Bethesda III–IV cytology and no surgical histology, and 20 public-dataset images excluded at the second verification pass because the region of interest did not meet the inclusion criteria or image quality was insufficient on double review) contributed neither to training nor to evaluation. Allocation to training, validation and test was performed a priori at patient level: the first 750-record block was assigned to training and the remaining patients were randomized with stratification on outcome (seed 42); the partition was locked with a SHA-256 checksum before any training step. The test set was never inspected and no hyperparameter was tuned on it. All 256 test nodules carried an independent EU-TIRADS rating, so the index test and the comparator were compared on an identical population. FNA = fine-needle aspiration, ROI = region of interest.

### Image acquisition and pre-processing

In the Tunisian centers, the EU-TIRADS category, the nodule measurements and the qualitative descriptors were taken from the routine radiology report of each examination, extracted from the radiology information systems. Images were exported as PNG or JPEG files from DICOM archives. The scanners of the Tunisian centers (Hitachi Ascendus and Fujifilm Arietta) and their high-frequency linear probes (> 7.5 MHz) were documented. Only B-mode images were given to the model; color and power Doppler images were not used as input. The central vascularity descriptor, assessed by the radiologist on color Doppler during the examination, entered Config D only as a text field of the structured report. Preprocessing comprised (i) selection of one representative image per nodule, linked to the clinical registry by a pseudonymized identifier; (ii) automated extraction of a bounding box around the region of interest with a 10% margin, followed by visual verification of each crop; (iii) quality control excluding images with more than 20% of the region of interest obscured; (iv) anonymization by masking burned-in metadata at the periphery of the image; and (v) normalization of intensities to the 0–1 range and resizing to 896 × 896 pixels, the native resolution of the MedSigLIP vision encoder [18]. No smoothing filter was applied, so that the native speckle texture was preserved.

### Model, fine-tuning and configurations

The base model was MedGemma 1.5 4B (google/medgemma-1.5-4b-it) [19], a medical vision–language model derived from Gemma 3 that pairs a MedSigLIP vision encoder (SigLIP ViT-So400m; 896 × 896 input, 14 × 14 patches) with a language decoder [18]. Tabular clinical and sonographic variables were written as structured natural-language text in the user prompt rather than passed through a separate branch of the network. This adds no parameters and allows inputs to be removed simply by editing the prompt (Figure 2).

**Figure 2.**
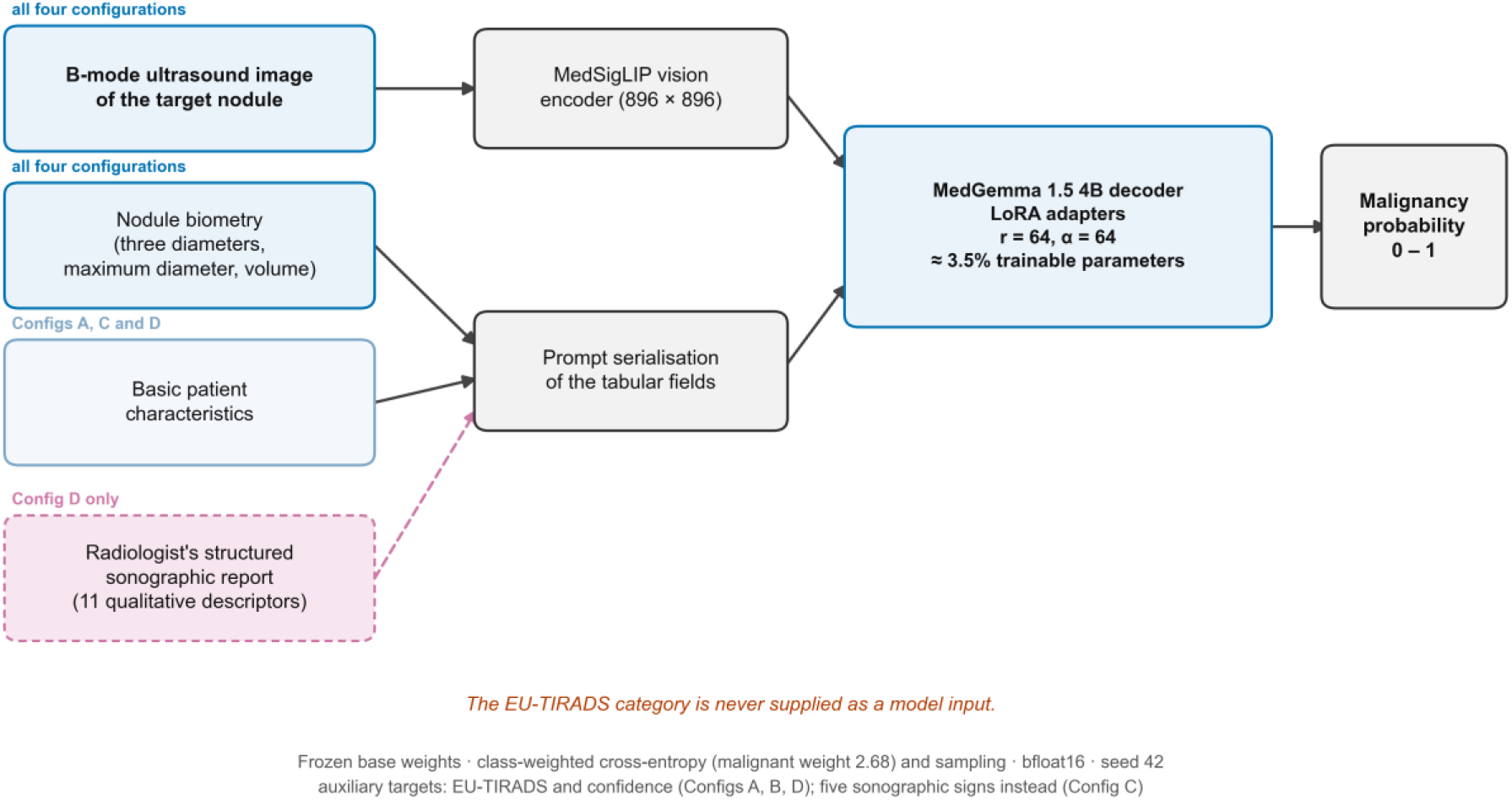
Overview of the multimodal pipeline. The B-mode image of the target nodule is encoded by MedSigLIP at its native 896 × 896 resolution; the fields of the structured report are serialized as natural-language text in the user prompt rather than passed through a separate architectural branch. The base weights of MedGemma 1.5 4B remain frozen; low-rank adapters on all linear layers (r = 64, α = 64) and the multimodal projector are trained (≈3.5% of parameters). The output is a continuous malignancy probability, to which a decision threshold is applied downstream. Training combines the primary malignancy target with auxiliary EU-TIRADS and confidence targets, replaced in Config C by five sonographic targets supervised by the recorded annotations. The overall EU-TIRADS category is never supplied as a model input.

Adaptation used LoRA [20] on all linear layers, with the multimodal projector trained in full and the base weights frozen (rank r = 64, α = 64, dropout 0.05), which left ≈157 million trainable parameters, ≈3.5% of the parameters of the adapted model (frozen base weights plus adapters). Optimization used AdamW (learning rate 2 × 10⁻⁴, weight decay 0.01, cosine schedule with 10% warm-up, 5% for Config C, gradient clipping at 1.0), an effective batch size of 16 (micro-batch of 4 with 4 accumulation steps), a sequence length of 768 tokens (Configs A and B) or 1,024 tokens (Configs C and D), and at most 10 epochs, with early stopping on validation performance evaluated every 50 optimization steps (patience of 5 evaluations). Class imbalance was handled by class-weighted cross-entropy (benign 0.615, malignant 2.677), combined with class-weighted sampling of training examples (malignant weight 5.0, about 1,630 examples per epoch in the training logs), rather than by synthetic oversampling, which creates artificial examples and can distort latent representations. The class weights were derived from the class frequencies of the analyzed cohort as n/(2 × n_class); the training partition alone gives nearly identical values (0.609 and 2.787). On-the-fly augmentation (probability 0.5 each) preserved native semiology: horizontal flip, photometric adjustment (± 15%), Gaussian blur (σ between 0.1 and 1.0) and affine rotation restricted to ± 10°; elastic deformation and vertical flip were not used. For Config D, each report field was masked at random during training (probability 0.10–0.15 per field), and all fields together in 10% of examples, to limit reliance on the text. Training ran locally in bfloat16 (CUDA 12.8) on an NVIDIA RTX PRO 6000 (Configs A and B; PyTorch 2.8.0) and an NVIDIA A100-SXM4 80 GB (Configs C and D; PyTorch 2.10.0), with a global random seed of 42.

Four configurations were carried forward to the sealed evaluation (Table 1). The configurations differ mainly in their inputs and auxiliary targets; the other differences are listed in Supplementary Tables S5–S6.

- **Config A:** image + nodule biometry (three orthogonal diameters, maximum diameter, volume) together with age and sex.
- **Config B:** image + nodule biometry alone.
- **Config C:** the inputs of Config A, with corrected logit extraction and chain-of-thought prompting whose five intermediate steps (echogenicity, margins, halo, microcalcifications, shape) are supervised by the recorded annotations, and without the auxiliary EU-TIRADS learning target; this is the prespecified sensitivity analysis bounding co-supervision bias.
- **Config D:** image + a 16-field structured prompt, made of the five prompt fields of Config A (age; sex; nodule volume; dimensions, that is, the three orthogonal and the maximum diameters; and size-derived ratios) and the eleven qualitative sonographic descriptors recorded by the radiologist (composition, spongiform appearance, echogenicity, shape, margins, microcalcifications, macrocalcifications, perinodular halo, central vascularity read on color Doppler, capsular contact and echogenic foci).

**Table 1.** The four evaluated model configurations.

| Configuration | Inputs in addition to the B-mode image | Auxiliary<br>EU-TIRADS<br>learning<br>target | Rationale |
| --- | --- | --- | --- |
| Config A | Nodule biometry (three orthogonal diameters, maximum diameter, volume) + age and sex | Yes | Reference multimodal configuration; no radiologist input beyond the image |
| Config B | Nodule biometry alone (three orthogonal diameters, maximum diameter, volume) | Yes | Input-restricted variant isolating nodule biometry; no radiologist input beyond the image |
| Config C | As Config A, with corrected logit extraction and chain-of-thought prompting supervised by the recorded annotations (echogenicity, margins, halo, microcalcifications, shape) | No | Prespecified sensitivity analysis bounding co-supervision bias; the five sonographic steps replace the EU-TIRADS auxiliary target; no qualitative radiologist interpretation |
| Config D | The inputs of Config A plus the eleven qualitative descriptors | Yes | “Radiologist assistant”: |
|  | of the structured radiology report (composition, spongiform appearance, echogenicity, shape, margins, microcalcifications, macrocalcifications, perinodular halo, central vascularity read on color Doppler, capsular contact, echogenic foci), 16 input fields in all |  | standardizes the report-to-risk mapping |

Configs A, B and C therefore need no qualitative interpretation from the radiologist: they work on the ultrasound image and routinely measured nodule dimensions, with or without the basic patient characteristics available on any request form. Config D is the only configuration that uses the radiologist’s structured interpretation. The exact variable list and training settings of each configuration are given in Supplementary Tables S4–S6.

Configs A, B and D were trained on several tasks at once, combining the primary malignancy target with an auxiliary EU-TIRADS target and an auxiliary confidence target; Config C replaced the EU-TIRADS target with five auxiliary sonographic targets supervised by the recorded annotations. The overall EU-TIRADS category was never given to any configuration as an input, which keeps the human comparator independent; its effect as an auxiliary learning target in three of the four configurations is bounded by Config C, which omits it. The scanner model, the free-text cytology report and the Bethesda class were also withheld, the last because it constitutes the label.

### Data partition and sealing

The cohort was partitioned a priori at patient level, so that no patient appeared in two partitions, in two steps. Patients with any nodule among the first 750 records of the database, all from the Tunis center and examined in 2019–2022, were assigned to training; the remaining patients were allocated to training, validation and test by outcome-stratified random sampling (seed 42), with a target of at least 95 malignant nodules in validation and test combined. The partition was versioned, locked with a SHA-256 checksum and applied identically in every analysis. The 256 test nodules were sealed before any training procedure, and neither visual inspection of this subset nor hyperparameter tuning on it was allowed. All 256 test nodules carried an independent EU-TIRADS rating, so the comparison was strictly paired on an identical population. No imputation of missing structured fields was planned; their completeness is reported in the Results. The database records only the year of each examination, so the interval between the index image, FNA and, when applicable, surgery could not be determined.

### Sample size

The required sample size was derived from Buderer’s formula for diagnostic studies [38], assuming a sensitivity of 0.90, a specificity of 0.75, precision margins of 0.05 and 0.07, and a malignancy prevalence of 0.12. In the absence of exhaustive Tunisian data, this prevalence was taken from recent series in the Middle East and North Africa (10.1% in the United Arab Emirates [39] and 13.8% in Yemen [40]). The calculation gave 139 malignant and 147 benign nodules, a total of ≈1,158 nodules, raised to ≈1,390 with a 20% safety margin. It sizes the overall cohort; because accuracy is estimated on the smaller sealed test set, the half-width of the sensitivity interval actually achieved is necessarily wider than the a priori target. The comparison of two correlated ROC curves follows Obuchowski’s method [41]; the precision achieved on the paired difference, its standard error and confidence interval, is reported in the Results and read against the prespecified margin of 0.05.

### Hypotheses

The primary hypothesis, fixed before evaluation, was that the fine-tuned model would reach an AUROC of at least 0.89 on the sealed test set. The hypothesis concerned the fine-tuned model, and results are reported for all four configurations. This threshold was set deliberately below the summary AUROC of 0.92–0.93 reported by recent meta-analyses of AI in thyroid ultrasound [12,13], to leave a statistical safety margin, while remaining more demanding than the threshold of 0.80 used in early deep learning studies. For the comparison with EU-TIRADS, a non-inferiority margin of 0.05 in AUROC was prespecified.

### Statistical analysis

All analyses were performed in Python 3.12 (scikit-learn, statsmodels and SciPy) with a global seed of 42, and two-sided p < 0.05 was considered significant. Every test-set value was recomputed from the locked prediction file, and the complete analysis was reproduced with Python 3.13 (scikit-learn 1.8.0 and SciPy 1.17.1) with identical results; validation-set values come from the training logs.

Continuous variables are summarized by median and interquartile range and compared with the Mann– Whitney U test; categorical variables are given as counts and percentages and compared with Pearson’s χ² test (Fisher’s exact test when expected counts were < 5). Proportions, including malignancy prevalence, are reported with Wilson score intervals [42].

The prespecified comparator was EU-TIRADS dichotomized at ≥ 4, a risk classifier that must be distinguished from the clinical indication for FNA, which also takes nodule size into account; the ≥ 5 (rule-in) threshold is reported as a sensitivity analysis. For threshold-independent comparison, EU-TIRADS categories were mapped to working probabilities fixed by the authors before the analysis (T2 → 0.02, T3 → 0.07, T4 → 0.20, T5 → 0.55): the value for category 5 follows the mean risk reported by the European consensus for nodules with high-risk features [3], the values for categories 3 and 4 lie within the 95% confidence intervals of the pooled malignancy prevalences of a meta-analysis of EU-TIRADS [43], and category 2 corresponds to a risk close to zero. These are not published point estimates. Because AUROC is invariant to monotone transformation, this mapping does not affect discrimination; it affects only the Brier score, the calibration metrics and the net benefit of EU-TIRADS, and the coincidence of the 20% decision-curve threshold with the probability assigned to category 4 follows from it. Config D was also compared with EU-TIRADS ≥ 5 by McNemar’s exact test on correct classifications, and the size-adapted EU-TIRADS indication for FNA (category 3 > 20 mm, 4 > 15 mm, 5 > 10 mm) was evaluated as a classifier.

The EU-TIRADS-anchored decision guide proposed in Figure 7 was applied to the sealed test set. EU-TIRADS 2–3 nodules kept their usual management, Config D serving only as a safety net, and EU-TIRADS 5 nodules kept confirmatory FNA. For an EU-TIRADS 4 nodule, FNA was deferred when Config D returned a benign output and no EU-TIRADS high-risk descriptor (marked hypoechogenicity, angular or lobulated margins, microcalcifications or a taller-than-wide shape) was present.

Discrimination is reported as AUROC and area under the precision–recall curve (AUPRC) [44], with 95% bias-corrected and accelerated (BCa) bootstrap intervals (B = 2,000) [45]. Classification metrics (sensitivity, specificity, predictive values, likelihood ratios, F1 and Matthews correlation coefficient) are reported at three operating points: the native deployment threshold of the model, applied blind; the Youden threshold derived on the validation partition alone (0.335), the only one fixed without access to the test data; and the Youden threshold recalculated on the test set, an optimistic upper bound kept as the prespecified point of comparison with EU-TIRADS. Config D was also evaluated across a grid of probability thresholds from 0.05 to 0.60, to describe the trade-off between cancers detected and aspirations indicated. Calibration is reported as Brier score, Brier skill score against a predictor using the prevalence alone, expected calibration error (ECE, 10 equal-width bins), calibration-in-the-large, Cox calibration slope and Spiegelhalter’s z test [46]. Because the raw softmax output was expected to be miscalibrated [47], Platt recalibration was estimated by internal 5-fold cross-validation with out-of-fold predictions. Clinical utility is expressed as net benefit in decision curve analysis [48,49]; a nodule was counted positive when its probability was greater than or equal to the threshold.

The model–comparator comparison used three prespecified methods on the sealed test set: DeLong’s test for correlated AUROCs [50]; binary net reclassification improvement (NRI) against EU-TIRADS ≥ 4, counting movement in both directions, with percentile bootstrap intervals (B = 10,000) [51]; and comparative decision curve analysis. Agreement between binary classifications was quantified by Cohen’s κ and McNemar’s test with continuity correction. Prespecified subgroup analyses covered sex, age, center, nodule size and EU-TIRADS category; strata with fewer than 10 events or 10 non-events are flagged as not interpretable. Holm correction was applied to the family of four comparisons with EU-TIRADS; p values for comparisons between configurations are unadjusted and are interpreted accordingly.

## Results

### Cohort

Of 1,580 documented nodules, 1,456 met the reference-standard criterion (272 malignant; prevalence 18.7%, 95% CI 16.8–20.8) and were allocated to training (942; 169 malignant, 17.9%), validation (258; 52 malignant, 20.2%) and the sealed test set (256; 51 malignant, 19.9%, 95% CI 15.5–25.2). These 1,456 nodules came from 1,280 patients (training 808, validation 236, test 236). The 124 excluded nodules comprised 104 Tunisian nodules with Bethesda III–IV cytology and no surgical histology (Tunis 96, Mahdia 8) and 20 public-dataset images excluded at the second verification pass (region of interest not meeting the inclusion criteria, or insufficient quality on double review) (Figure 1). The reference standard was cytological for 1,342 Tunisian nodules and histological for 114: 11 Tunisian nodules, all malignant (Bethesda II, III and IV: 1, 3 and 7), and the 103 public-dataset nodules. The 16 structured input fields were complete for all 1,456 included nodules, so no imputation was needed. The Tunis center contributed 1,233 nodules (84.7%) to the cohort and 168 (65.6%) to the test set.

The cohort was predominantly female (1,260/1,456; 86.5%) with a median age of 51 years (IQR 39–62). Patients with malignancy were younger (median 46 vs 53 years; p < 0.001). Malignancy prevalence did not differ across the three sources (Mahdia 16.7%, public dataset 19.4%, Tunis 18.8%; p = 0.83), nor by sex (19.4% in men vs 18.6% in women; p = 0.86). EU-TIRADS categories were distributed as 2 (0.5%), 3 (41.0%), 4 (38.6%) and 5 (19.8%); malignancy prevalence rose monotonically across categories (0%, 1.2%, 6.8%, 78.5%; Cochran–Armitage p < 0.001), and 97.4% of cancers (265/272) were in categories 4– 5 (Supplementary Figure S1). Malignant nodules were smaller than benign ones (median maximum diameter 11 mm, IQR 8–15, vs 23 mm, IQR 18–28.2; p < 0.001; median volume 0.4, IQR 0.2–1.0, vs 2.7, IQR 1.2–5.4 cm³). Malignancy prevalence increased with the number of concurrent high-suspicion features, from 2.6% (29/1,114) with none to 100% (6/6) with all five (the four EU-TIRADS high-suspicion signs and capsular contact; Supplementary Figure S2). Full cohort characteristics are given in Table 2.

**Table 2.**
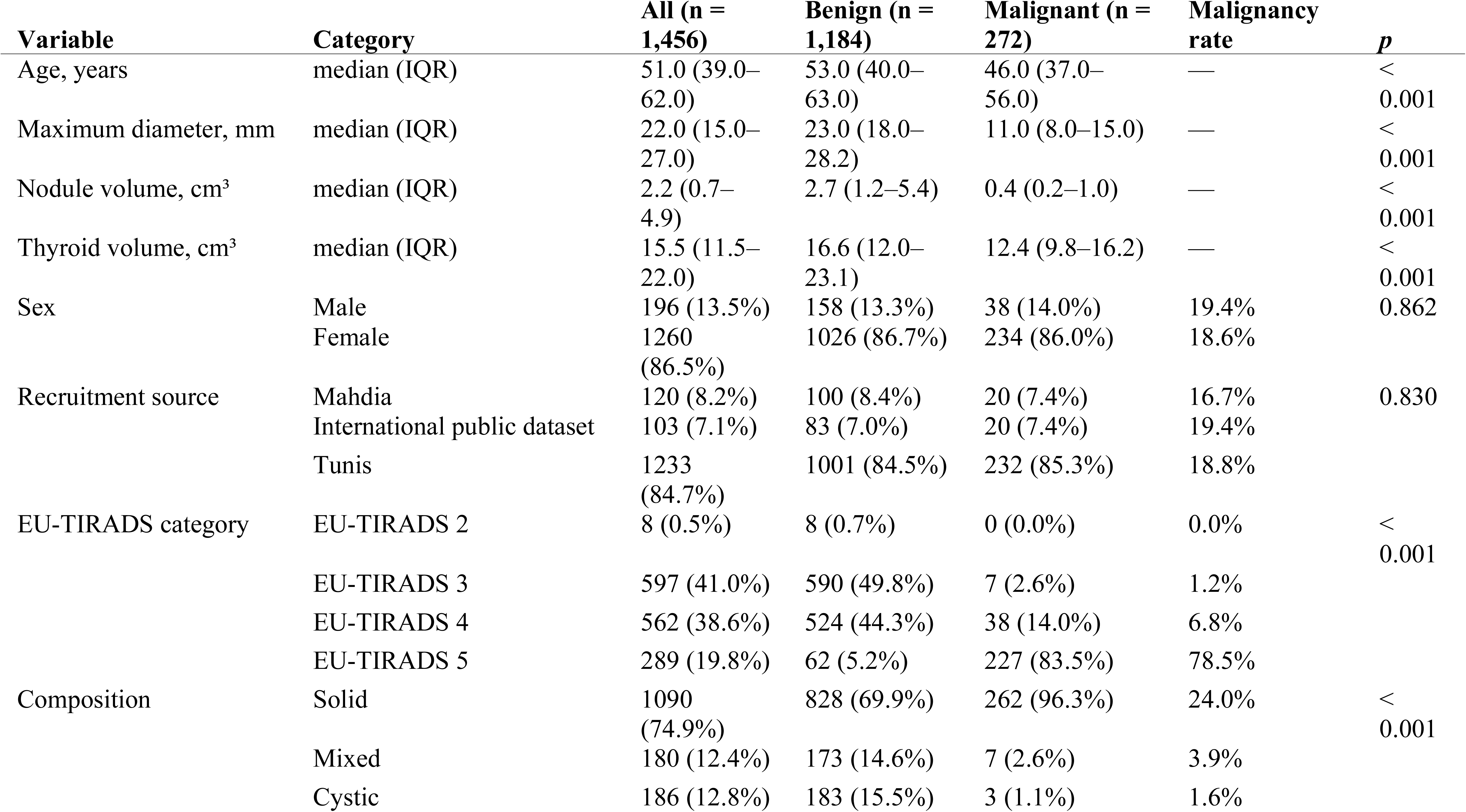
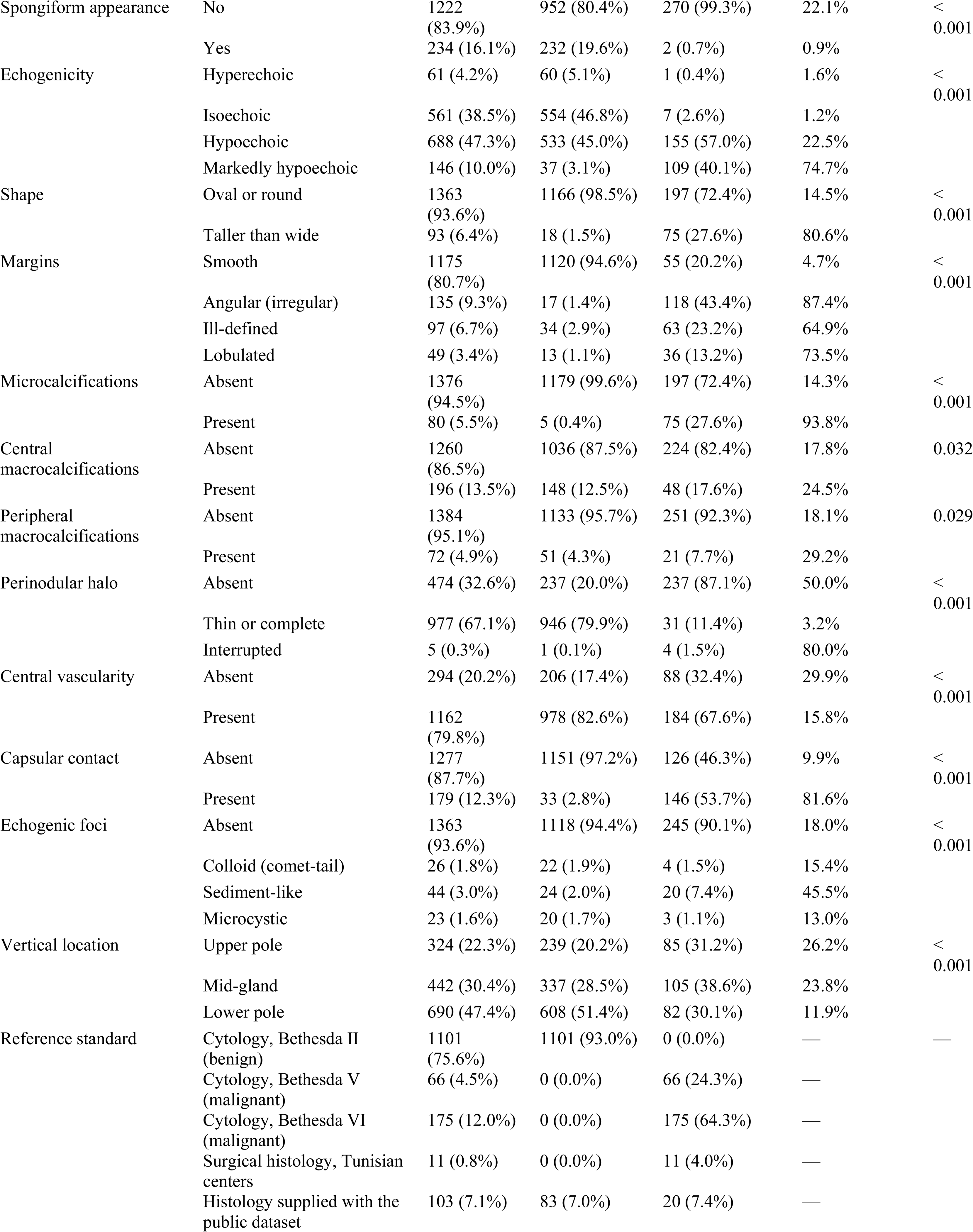
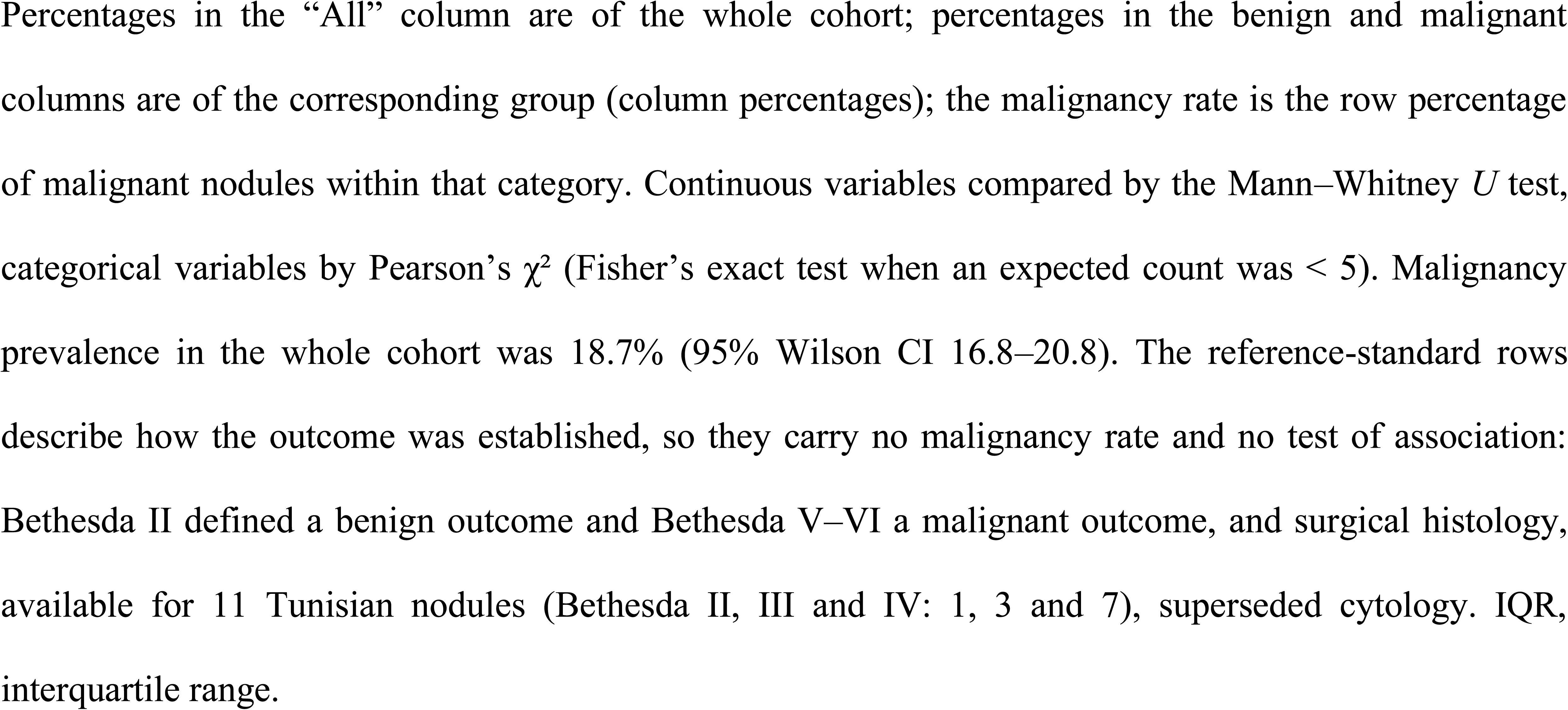
Characteristics of the analyzed cohort (n = 1,456 nodules)

### Discrimination on the sealed test set

Config D achieved the highest discrimination (AUROC 0.953, 95% CI 0.909–0.976; AUPRC 0.834, 95% CI 0.686–0.914). Configs A, B and C clustered near the prespecified threshold (AUROC 0.892, 0.886 and 0.883) with lower AUPRC (0.634, 0.712 and 0.627, respectively). Expert-rated EU-TIRADS reached an AUROC of 0.948 (0.894–0.972) and an AUPRC of 0.784 (0.664–0.876) on the same nodules (Figure 3, Table 3). The primary hypothesis (AUROC ≥ 0.89) was met by Config D, whose lower confidence bound (0.909) exceeded 0.89; Config A reached the threshold as a point estimate only (lower bound 0.841).

**Figure 3.**
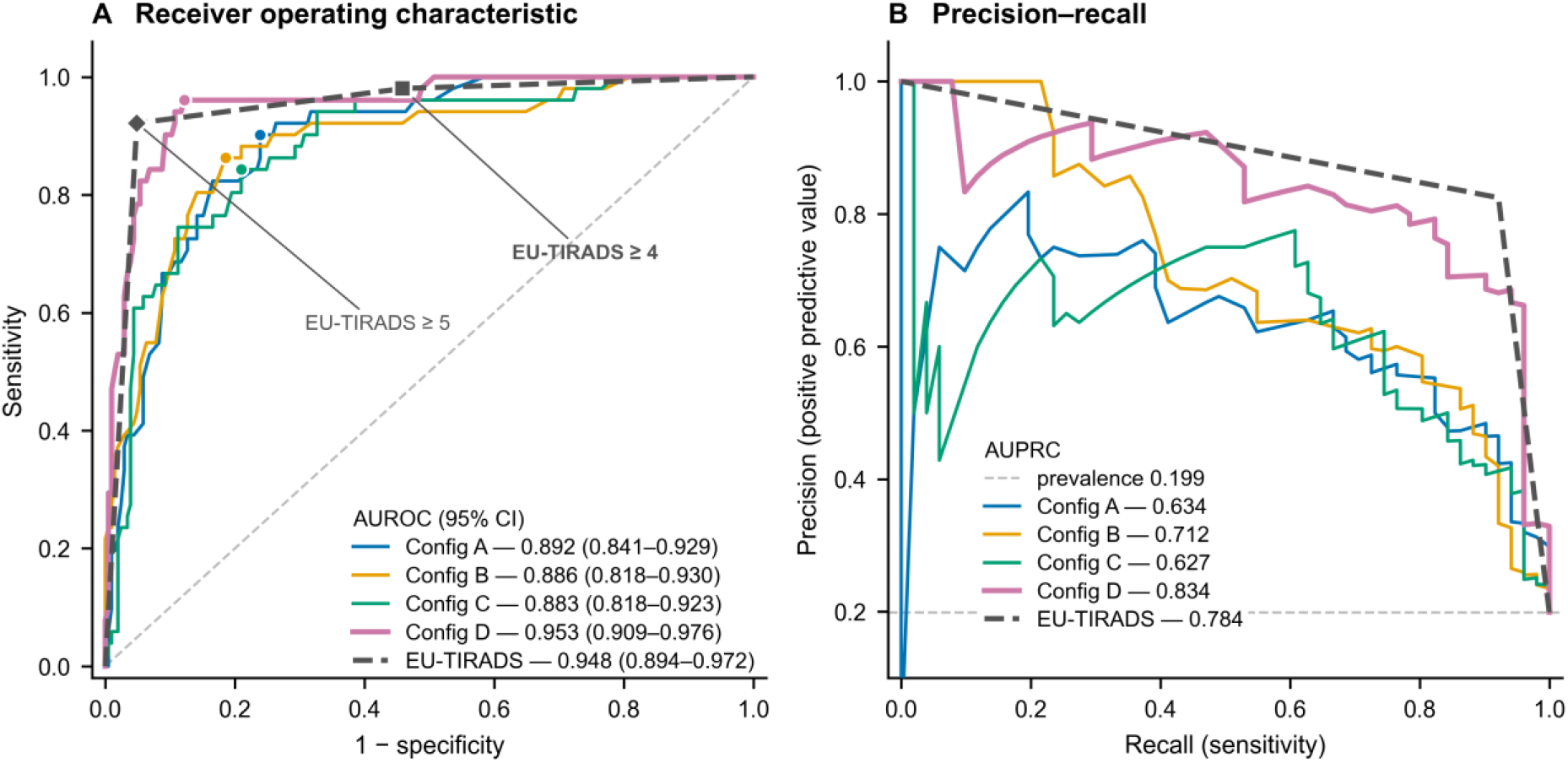
Discrimination on the sealed test set (n = 256; 51 malignant). (A) Receiver operating characteristic curves for the four fine-tuned configurations and for expert-rated EU-TIRADS, with areas under the curve and 95% bias-corrected and accelerated bootstrap intervals (B = 2,000, seed 42). Filled circles mark the Youden point of each model; the square and the diamond mark the EU-TIRADS ≥ 4 and ≥ 5 operating points. Config D and EU-TIRADS overlap closely (Δ AUROC +0.004; DeLong p = 0.66), whereas Configs A, B and C lie significantly below the score. (B) Precision–recall curves with areas under the curve; the dashed horizontal line is the test-set prevalence (0.199), the no-skill reference for this metric. The precision–recall view separates the configurations more sharply than the receiver operating characteristic view. AUPRC = area under the precision–recall curve, AUROC = area under the receiver operating characteristic curve.

**Table 3.** Diagnostic performance on the sealed test set (n = 256; 51 malignant, prevalence 19.9%)

| Metric | Config A | Config B | Config C | Config D | EU-TIRADS ≥ 4 | EU-TIRADS ≥ 5 |
| --- | --- | --- | --- | --- | --- | --- |
| **Threshold-independent discrimination** |  |  |  |  |  |  |
| AUROC | 0.892<br>(0.841–0.929) | 0.886<br>(0.818–0.930) | 0.883<br>(0.818–0.923) | 0.953<br>(0.909–0.976) | 0.948<br>(0.894–0.972) |  |
| AUPRC | 0.634<br>(0.459–0.750) | 0.712<br>(0.575–0.810) | 0.627<br>(0.464–0.746) | 0.834<br>(0.686–0.914) | 0.784<br>(0.664–0.876) |  |
| **Youden threshold of the test set (models); category threshold (EU-TIRADS)** |  |  |  |  |  |  |
| Decision threshold | 0.0026 | 0.0032 | 0.0003 | 0.3208 | category ≥ 4 | category ≥ 5 |
| TP / FN | 46 / 5 | 44 / 7 | 43 / 8 | 49 / 2 | 50 / 1 | 47 / 4 |
| TN / FP | 156 / 49 | 167 / 38 | 162 / 43 | 180 / 25 | 111 / 94 | 195 / 10 |
| Sensitivity, % | 90.2<br>(79.0–95.7) | 86.3<br>(74.3–93.2) | 84.3<br>(72.0–91.8) | 96.1<br>(86.8–98.9) | 98.0<br>(89.7–99.7) | 92.2<br>(81.5–96.9) |
| Specificity, % | 76.1<br>(69.8–81.4) | 81.5<br>(75.6–86.2) | 79.0<br>(72.9–84.0) | 87.8<br>(82.6–91.6) | 54.1<br>(47.3–60.8) | 95.1<br>(91.3–97.3) |
| PPV, % | 48.4<br>(38.6–58.3) | 53.7<br>(42.9–64.0) | 50.0<br>(39.7–60.3) | 66.2<br>(54.9–76.0) | 34.7<br>(27.4–42.8) | 82.5<br>(70.6–90.2) |
| NPV, % | 96.9<br>(92.9–98.7) | 96.0<br>(91.9–98.0) | 95.3<br>(91.0–97.6) | 98.9<br>(96.1–99.7) | 99.1<br>(95.1–99.8) | 98.0<br>(94.9–99.2) |
| LR+ / LR– | 3.77 / 0.129 | 4.65 / 0.168 | 4.02 / 0.198 | 7.88 / 0.045 | 2.14 / 0.036 | 18.89 / 0.082 |
| F1 / MCC | 0.630 / 0.548 | 0.662 / 0.580 | 0.628 / 0.536 | 0.784 / 0.739 | 0.513 / 0.420 | 0.870 / 0.838 |
| **Youden threshold of the validation partition (models); category threshold (EU-TIRADS)** |  |  |  |  |  |  |
| Decision threshold | 0.0014 | 0.0063 | 0.0005 | 0.3346 | category ≥ 4 | category ≥ 5 |
| TP / FN | 47 / 4 | 34 / 17 | 38 / 13 | 48 / 3 | 50 / 1 | 47 / 4 |
| TN / FP | 147 / 58 | 185 / 20 | 182 / 23 | 183 / 22 | 111 / 94 | 195 / 10 |
| Sensitivity, % | 92.2<br>(81.5–96.9) | 66.7<br>(53.0–78.0) | 74.5<br>(61.1–84.5) | 94.1<br>(84.1–98.0) | 98.0<br>(89.7–99.7) | 92.2<br>(81.5–96.9) |
| Specificity, % | 71.7<br>(65.2–77.4) | 90.2<br>(85.4–93.6) | 88.8<br>(83.7–92.4) | 89.3<br>(84.3–92.8) | 54.1<br>(47.3–60.8) | 95.1<br>(91.3–97.3) |
| PPV, % | 44.8<br>(35.6–54.3) | 63.0<br>(49.6–74.6) | 62.3<br>(49.7–73.4) | 68.6<br>(57.0–78.2) | 34.7<br>(27.4–42.8) | 82.5<br>(70.6–90.2) |
| NPV, % | 97.4<br>(93.4–99.0) | 91.6<br>(86.9–94.7) | 93.3<br>(88.9–96.1) | 98.4<br>(95.4–99.4) | 99.1<br>(95.1–99.8) | 98.0<br>(94.9–99.2) |
| LR+ / LR– | 3.26 / 0.109 | 6.83 / 0.369 | 6.64 / 0.287 | 8.77 / 0.066 | 2.14 / 0.036 | 18.89 / 0.082 |
| F1 / MCC | 0.603 / 0.519 | 0.648 / 0.557 | 0.679 / 0.593 | 0.793 / 0.747 | 0.513 / 0.420 | 0.870 / 0.838 |
| **Native output of the model, applied blind (models); category threshold (EU-TIRADS)** |  |  |  |  |  |  |
| Decision threshold | native output | native output | native output | native output | category ≥ 4 | category ≥ 5 |
| TP / FN | 34 / 17 | 29 / 22 | 31 / 20 | 45 / 6 | 50 / 1 | 47 / 4 |
| TN / FP | 184 / 21 | 197 / 8 | 194 / 11 | 193 / 12 | 111 / 94 | 195 / 10 |
| Sensitivity, % | 66.7<br>(53.0–78.0) | 56.9<br>(43.3–69.5) | 60.8<br>(47.1–73.0) | 88.2<br>(76.6–94.5) | 98.0<br>(89.7–99.7) | 92.2<br>(81.5–96.9) |
| Specificity, % | 89.8<br>(84.8–93.2) | 96.1<br>(92.5–98.0) | 94.6<br>(90.6–97.0) | 94.1<br>(90.0–96.6) | 54.1<br>(47.3–60.8) | 95.1<br>(91.3–97.3) |
| PPV, % | 61.8<br>(48.6–73.5) | 78.4<br>(62.8–88.6) | 73.8<br>(58.9–84.7) | 78.9<br>(66.7–87.5) | 34.7<br>(27.4–42.8) | 82.5<br>(70.6–90.2) |
| NPV, % | 91.5<br>(86.9–94.7) | 90.0<br>(85.3–93.3) | 90.7<br>(86.0–93.9) | 97.0<br>(93.6–98.6) | 99.1<br>(95.1–99.8) | 98.0<br>(94.9–99.2) |
| LR+ / LR– | 6.51 / 0.371 | 14.57 / 0.449 | 11.33 / 0.414 | 15.07 / 0.125 | 2.14 / 0.036 | 18.89 / 0.082 |
| F1 / MCC | 0.642 / 0.549 | 0.659 / 0.602 | 0.667 / 0.598 | 0.833 / 0.791 | 0.513 / 0.420 | 0.870 / 0.838 |

Values in parentheses are 95% confidence intervals. AUROC and AUPRC intervals are 95% bias-corrected and accelerated bootstrap intervals ( $B = 2,000$ , seed 42); intervals for proportions are Wilson score intervals. The models are reported at three operating points: the Youden threshold of the test set, an optimistic upper bound kept as the prespecified point of comparison with EU-TIRADS; the Youden threshold of the validation partition, fixed without access to the test data; and the native output of the model, applied blind. EU-TIRADS has no model threshold: its two category thresholds are repeated in each block for side-by-side comparison, and its AUROC and AUPRC, computed on the score itself, are common to both. TP, true positives; FN, false negatives; TN, true negatives; FP, false positives; PPV and NPV, positive and negative predictive values; LR+ and LR–, positive and negative likelihood ratios; MCC, Matthews correlation coefficient.

Because each nodule contributed exactly one image, image-level and nodule-level AUROC and AUPRC are identical by construction. Validation-set discrimination at the selected checkpoint tracked the same ordering (Config D 0.908; Configs A, B, C 0.817, 0.803, 0.779), with best checkpoints between epochs 2 and 4 (Supplementary Table S7).

### Formal comparison with EU-TIRADS

By DeLong’s test, Configs A (Δ AUROC −0.056; p = 0.015), B (−0.062; p = 0.011) and C (−0.066; p = 0.018) were significantly inferior to EU-TIRADS ≥ 4 (Holm-adjusted p = 0.045 for each). Config D was not distinguishable from it (Δ +0.004, 95% bootstrap CI −0.016 to +0.025; p = 0.66); the lower bound of that interval (−0.016) lay above the prespecified non-inferiority margin of −0.05, so the prespecified non-inferiority criterion was met. Within the model family, Config D was superior to Config A (Δ +0.061; p = 0.012), Config B (+0.067; p = 0.015) and Config C (+0.070; p = 0.012), whereas Configs A, B and C were statistically indistinguishable from one another (all p > 0.59). This includes Config C, which removes the auxiliary EU-TIRADS target and differed from Config A by 0.009 AUROC (Table 4).

**Table 4.** Formal comparison of AUROCs (DeLong test for correlated curves, sealed test set, n = 256)

| Comparison | AUROC, first | AUROC, second | $\Delta$ AUROC (first – second) | 95% CI of $\Delta$ | $z$ | $p$ |
| --- | --- | --- | --- | --- | --- | --- |
| *Versus the prespecified comparator, EU-TIRADS $\geq 4$* |  |  |  |  |  |  |
| Config A vs EU-TIRADS | 0.892 | 0.948 | –0.056 | –0.100 to –0.013 | –2.442 | 0.015 |
| Config B vs EU-TIRADS | 0.886 | 0.948 | –0.062 | –0.110 to –0.017 | –2.535 | 0.011 |
| Config C vs EU-TIRADS | 0.883 | 0.948 | –0.066 | –0.122 to –0.015 | –2.374 | 0.018 |
| Config D vs EU-TIRADS | 0.953 | 0.948 | +0.004 | –0.016 to +0.025 | +0.439 | 0.660 |
| *Between configurations* |  |  |  |  |  |  |
| Config A vs Config B | 0.892 | 0.886 | +0.006 | — | +0.311 | 0.756 |
| Config A vs Config C | 0.892 | 0.883 | +0.009 | — | +0.532 | 0.595 |
| Config B vs Config C | 0.886 | 0.883 | +0.004 | — | +0.199 | 0.842 |
| Config A vs Config D | 0.892 | 0.953 | –0.061 | — | –2.507 | 0.012 |

| Comparison | AUROC,<br>first | AUROC,<br>second | $\Delta$ AUROC (first –<br>second) | 95% CI of $\Delta$ | z | p |
| --- | --- | --- | --- | --- | --- | --- |
| Config B vs Config D | 0.886 | 0.953 | −0.067 | — | −2.431 | 0.015 |
| Config C vs Config D | 0.883 | 0.953 | −0.070 | — | −2.521 | 0.012 |

Two-sided DeLong test on correlated AUROCs (identical nodules for both classifiers). EU-TIRADS was mapped to a continuous score using working category probabilities fixed by the authors before the analysis, which are not published point estimates ( $T2 \rightarrow 0.02$ , $T3 \rightarrow 0.07$ , $T4 \rightarrow 0.20$ , $T5 \rightarrow 0.55$ ; Materials and Methods), and dichotomized at $\geq 4$ for the prespecified comparison; because AUROC is invariant to monotone transformation, the mapping does not affect these results. Confidence intervals for $\Delta$ AUROC are percentile bootstrap intervals ( $B = 2,000$ , seed 42). The prespecified non-inferiority margin was $\Delta$ AUROC = 0.05. p values are unadjusted; after Holm correction over the four comparisons with EU-TIRADS, Configs A, B and C remain inferior (adjusted p = 0.045 each) and Config D remains indistinguishable (p = 0.660).

The paired difference between Config D and EU-TIRADS was estimated with a standard error of 0.010 (95% CI −0.016 to +0.025); with this standard error, the smallest difference detectable with 80% power was 0.029.

### Operating points and reclassification

At the prespecified ≥ 4 threshold, EU-TIRADS detected 50 of 51 cancers (sensitivity 98.0%, 95% CI 89.7–99.7), with 94 false positives among 205 benign nodules (specificity 54.1%, 47.3–60.8; positive predictive value 34.7%; negative predictive value 99.1%; LR+ 2.14, LR− 0.036). At its Youden threshold, Config D reached a sensitivity of 96.1% (86.8–98.9) and a specificity of 87.8% (82.6–91.6), with a positive predictive value of 66.2% and a negative predictive value of 98.9% (LR+ 7.88, LR− 0.045). At the threshold derived on the validation partition alone (0.335), applied unchanged, it reached 94.1% sensitivity and 89.3% specificity (48 true positives, 183 true negatives, 22 false positives, 3 false negatives). At its native deployment threshold, applied blind, Config D gave 88.2% sensitivity and 94.1% specificity (45 true positives, 193 true negatives, 12 false positives, 6 false negatives; F1 0.833, MCC 0.791), the best F1 and MCC of the four configurations. At their native thresholds, the other configurations had a sensitivity of 56.9–66.7% and a specificity of 89.8–96.1% (Table 3, Figure 4A–B).

**Figure 4.**
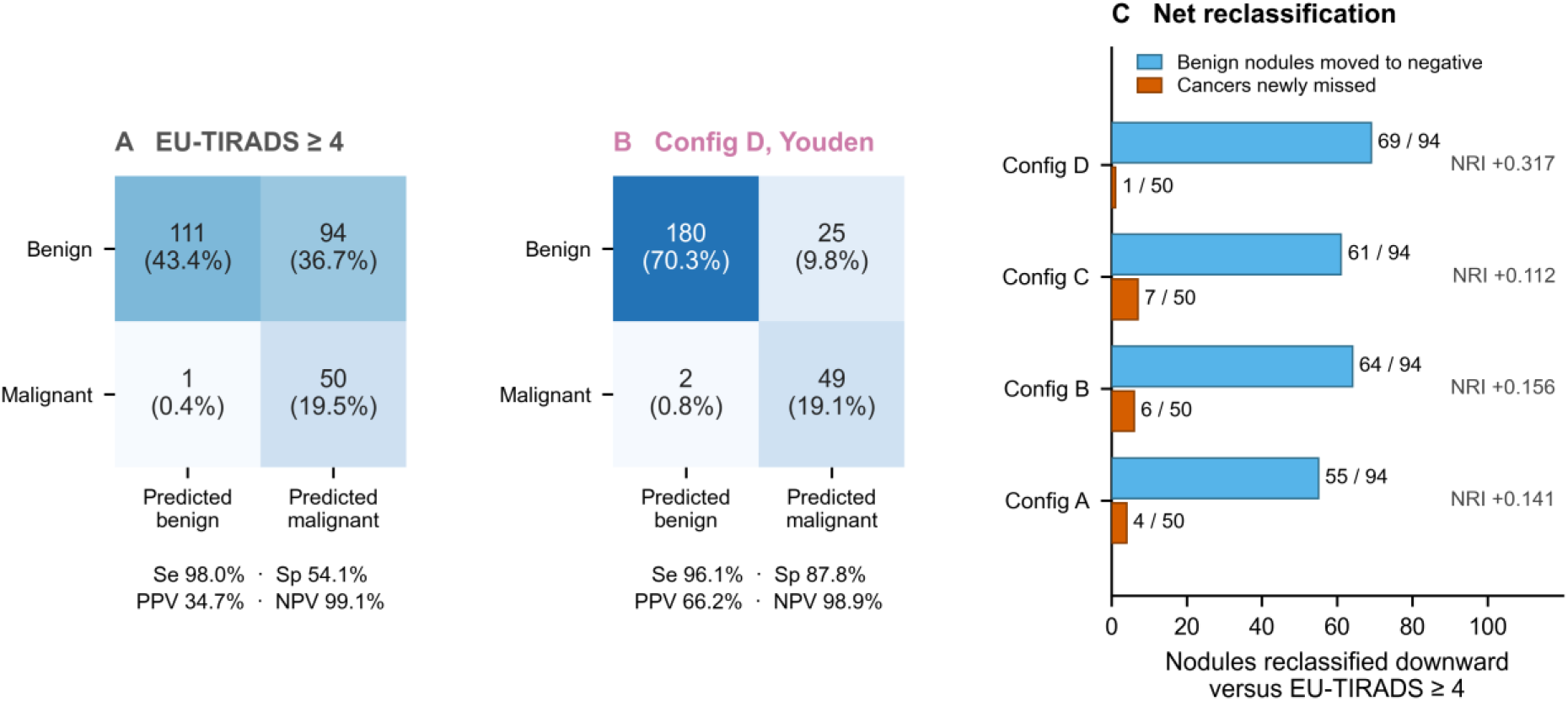
Operating points and reclassification against the comparator (sealed test set, n = 256). **(A)** Confusion matrix for EU-TIRADS at the prespecified ≥ 4 threshold: sensitivity 98.0%, with 94 false positives among 205 benign nodules. **(B)** Confusion matrix for Config D at its Youden threshold: sensitivity remains high (96.1%) while specificity rises to 87.8%. **(C)** Net downward reclassification of each configuration against EU-TIRADS ≥ 4. Blue bars count benign nodules called positive by the score and negative by the model (out of the 94 false positives the score produces); orange bars count cancers detected by the score and missed by the model (out of the 50 it detects). Config D reclassifies 69 benign nodules for a single newly missed cancer, giving the largest net reclassification improvement (+0.317). NRI values shown are the full binary net reclassification improvement, which also accounts for upward reclassification.

Against EU-TIRADS ≥ 4 at the Youden threshold, Config D moved 69 of the 94 false positives (73.4%) to negative while newly missing 1 of the 50 cancers EU-TIRADS had detected (2.0%), giving the largest net reclassification improvement of the four configurations (+0.317, 95% CI +0.240 to +0.390; p < 0.001; Table 5). Config D was also the only configuration that moved no benign nodule upward, so its raw and net benign gains coincided at 69. The other three moved 55, 64 and 61 benign nodules down but 10, 8 and 10 up, leaving net gains of 45, 56 and 51, and missed 4, 6 and 7 cancers the score had detected (NRI +0.141, +0.156 and +0.112) (Figure 4C, Table 5). The corresponding McNemar table for Config D versus EU-TIRADS ≥ 4 was asymmetric (69 discordances favoring the model versus 1 favoring the score; χ² = 64.1, p < 0.001). Broken down by category, the 94 false positives comprised 84 EU-TIRADS 4 and 10 EU-TIRADS 5 benign nodules; Config D reclassified 68 and 1 of them at the Youden threshold and 71 and 1 at the validation-derived threshold, and the one and two cancers it newly missed were among the three EU-TIRADS 4 cancers.

**Table 5.**
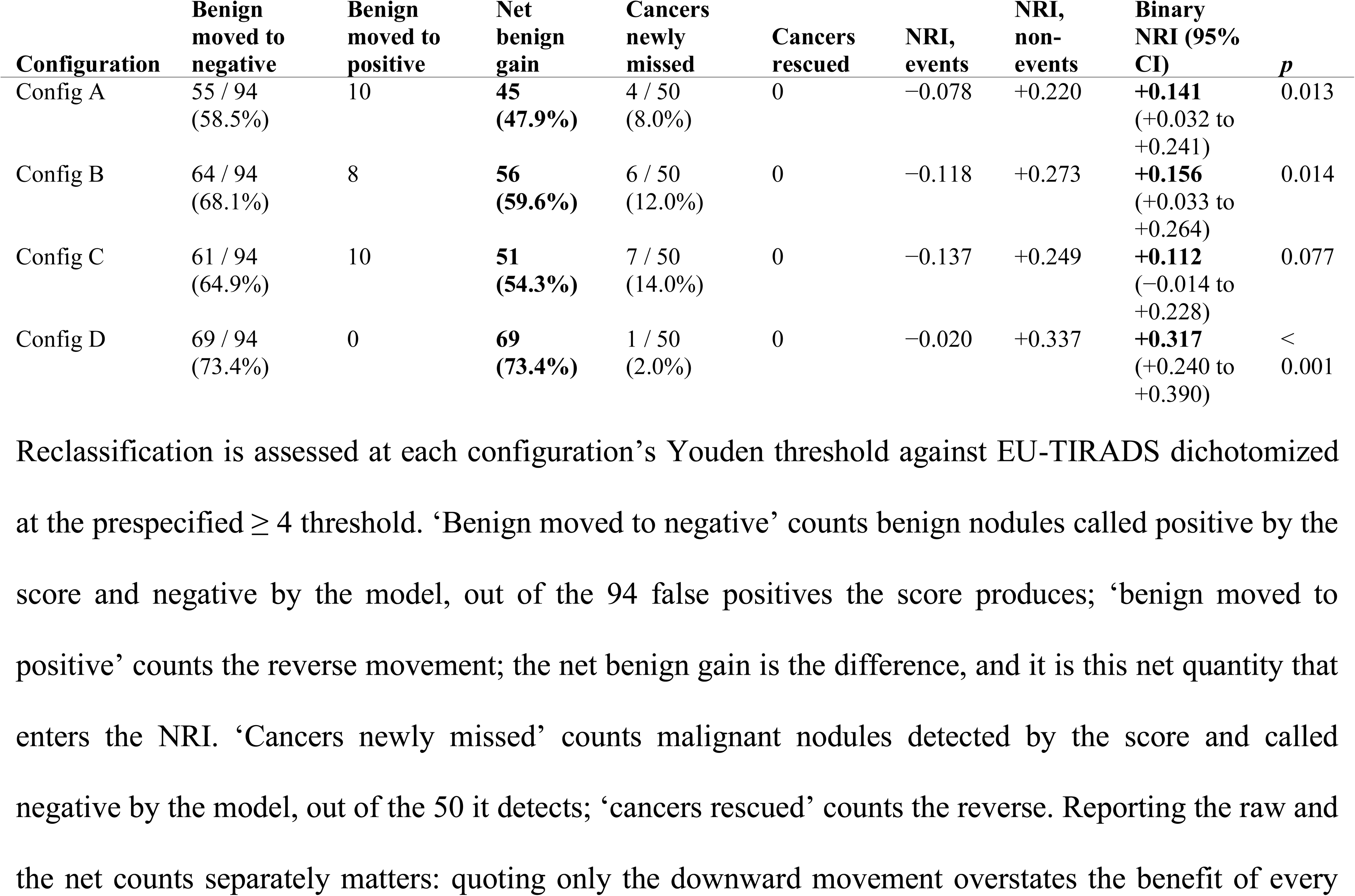
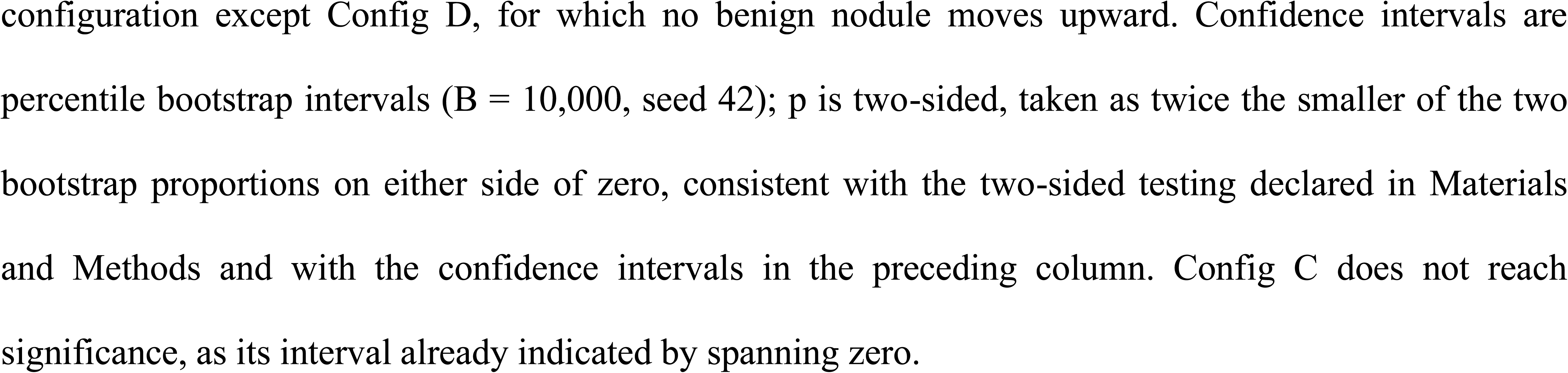
Net reclassification against EU-TIRADS ≥ 4 (sealed test set, n = 256)

Categorical agreement between the model’s binary output and EU-TIRADS depended on the threshold: at ≥ 4, κ was 0.218–0.364 across configurations (raw agreement 57.4–66.0%); at the high-suspicion threshold ≥ 5, agreement was higher, reaching κ = 0.932 (95% CI 0.872–0.985; raw agreement 97.7%) for Config D (Table 6). Against EU-TIRADS ≥ 5 (47 true positives, 4 false negatives, 10 false positives, 195 true negatives), Config D at its native threshold classified correctly one nodule that the ≥ 5 threshold misclassified and misclassified five that it classified correctly (McNemar exact p = 0.22); at the Youden and validation-derived thresholds, these counts were 3 versus 16 (p = 0.004) and 2 versus 13 (p = 0.007), Config D detecting two and one additional cancers at the cost of 15 and 12 additional false positives. Used as a classifier, the size-adapted EU-TIRADS indication for FNA had a sensitivity of 47.1% and a specificity of 16.1% on the sealed test set (24 of 51 cancers and 33 of 205 benign nodules correctly classified); the median maximum diameter of the test-set cancers was 11 mm.

**Table 6.** Agreement between the model’s binary output and EU-TIRADS (Cohen’s κ, sealed test set)

| Configuration | $\kappa$ vs EU-TIRADS $\geq 4$ (95% CI) | Raw agreement | $\kappa$ vs EU-TIRADS $\geq 5$ (95% CI) | Raw agreement |
| --- | --- | --- | --- | --- |
| Config A | 0.307 (0.222–0.389) | 62.9% | 0.657 (0.529–0.769) | 88.3% |
| Config B | 0.218 (0.151–0.291) | 57.4% | 0.587 (0.455–0.701) | 87.5% |
| Config C | 0.250 (0.176–0.320) | 59.4% | 0.689 (0.565–0.798) | 90.2% |
| Config D | 0.364 (0.283–0.445) | 66.0% | 0.932 (0.872–0.985) | 97.7% |

Model outputs binarized at the native deployment threshold. Intervals are percentile bootstrap intervals ( $B = 1,000$ , seed 42).

### Decision thresholds

The three operating points of Config D are the native output of the model, applied blind; the Youden threshold of the validation partition (0.335), the only threshold set without access to the test data; and the Youden threshold of the test set (0.321), an optimistic upper bound. Across thresholds, the exchange between cancers detected and aspirations indicated was steep at both ends and flat in between (Supplementary Table S8). At a threshold of 0.10, the model detected 50 of the 51 cancers, as EU-TIRADS ≥ 4 did, but with 100 false positives rather than 94. Between 0.15 and 0.321, the model detected 49 cancers while false positives fell from 72 to 25, and at 0.335 it detected 48 with 22 false positives. Above that range, each further reduction in aspirations cost cancers: 43 were detected at 0.40 (14 false positives) and 34 at 0.50 (7 false positives).

### Calibration

None of the four configurations was well calibrated in its raw form (Spiegelhalter p < 0.001 for all). Their failure modes differed: Configs A and B produced compressed probabilities that underestimated risk (Supplementary Figure S3; calibration-in-the-large +4.23 and +4.51; observed/expected ratios 29.4 and 43.2) and negative Brier skill scores (−0.200 and −0.214); Config C was intermediate (Brier skill score +0.295); Config D achieved the best Brier score of the models (0.080) and a Brier skill score of +0.496, close to EU-TIRADS (0.075 and +0.529), with a calibration-in-the-large of −0.28 (ideal 0) and a calibration slope of 2.51 (ideal 1) (Table 7, Figure 5A).

**Table 7.**
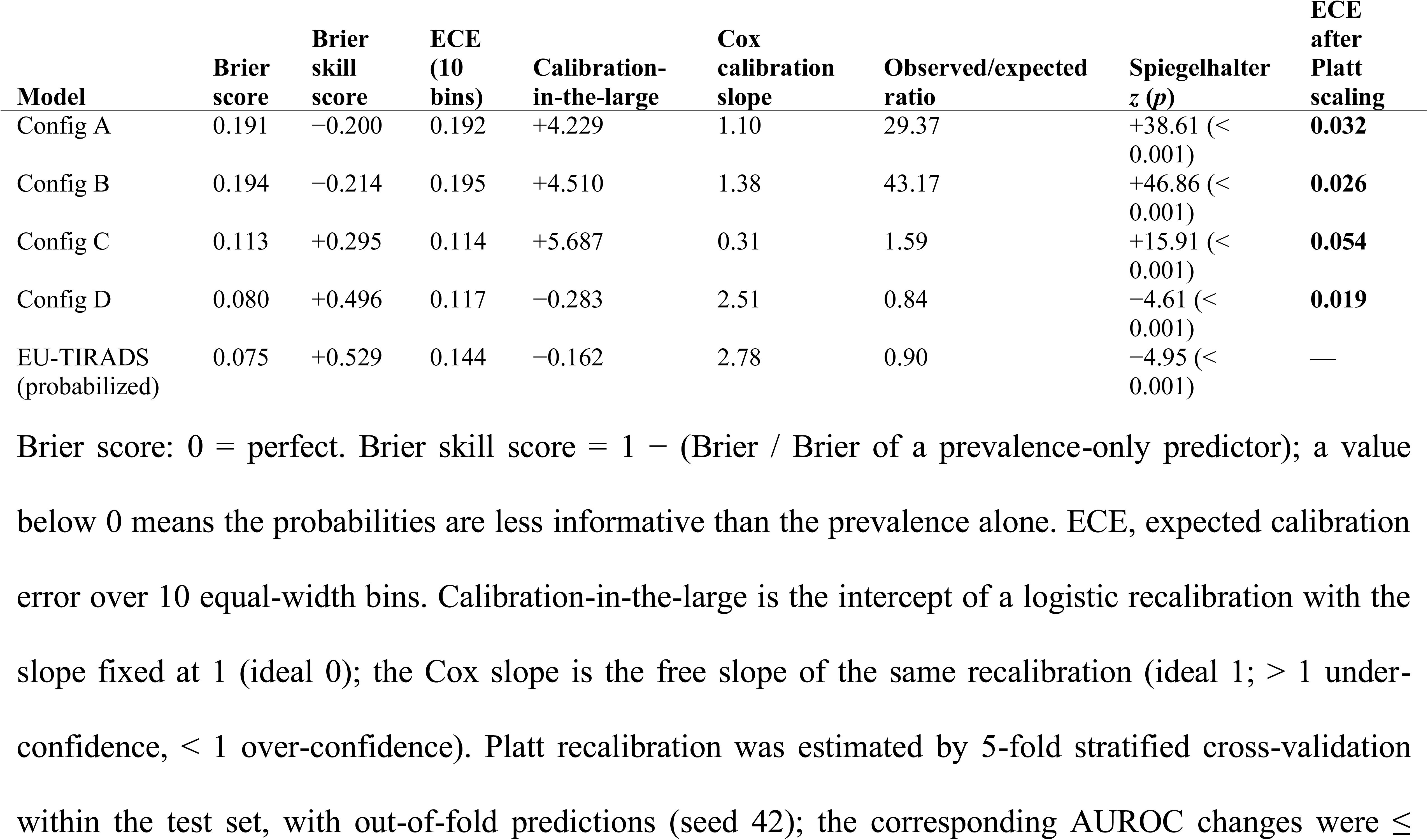
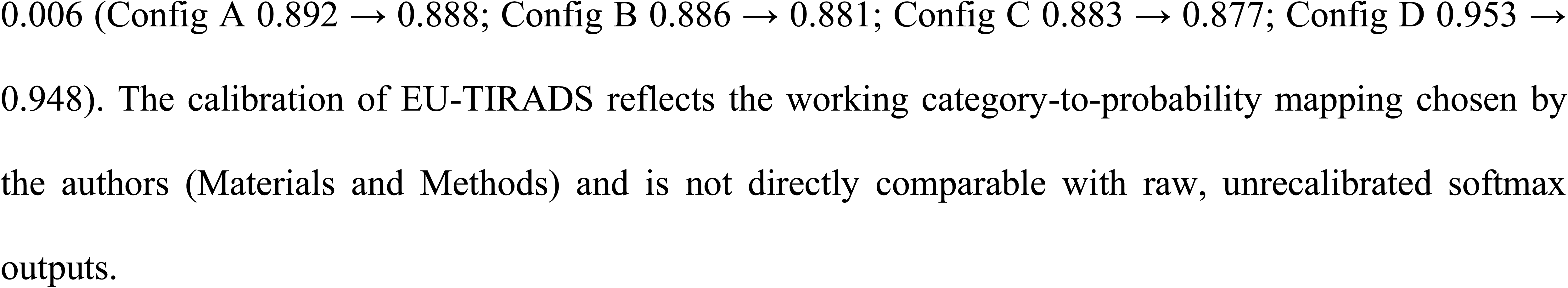
Probabilistic calibration on the sealed test set (n = 256)

**Figure 5.**
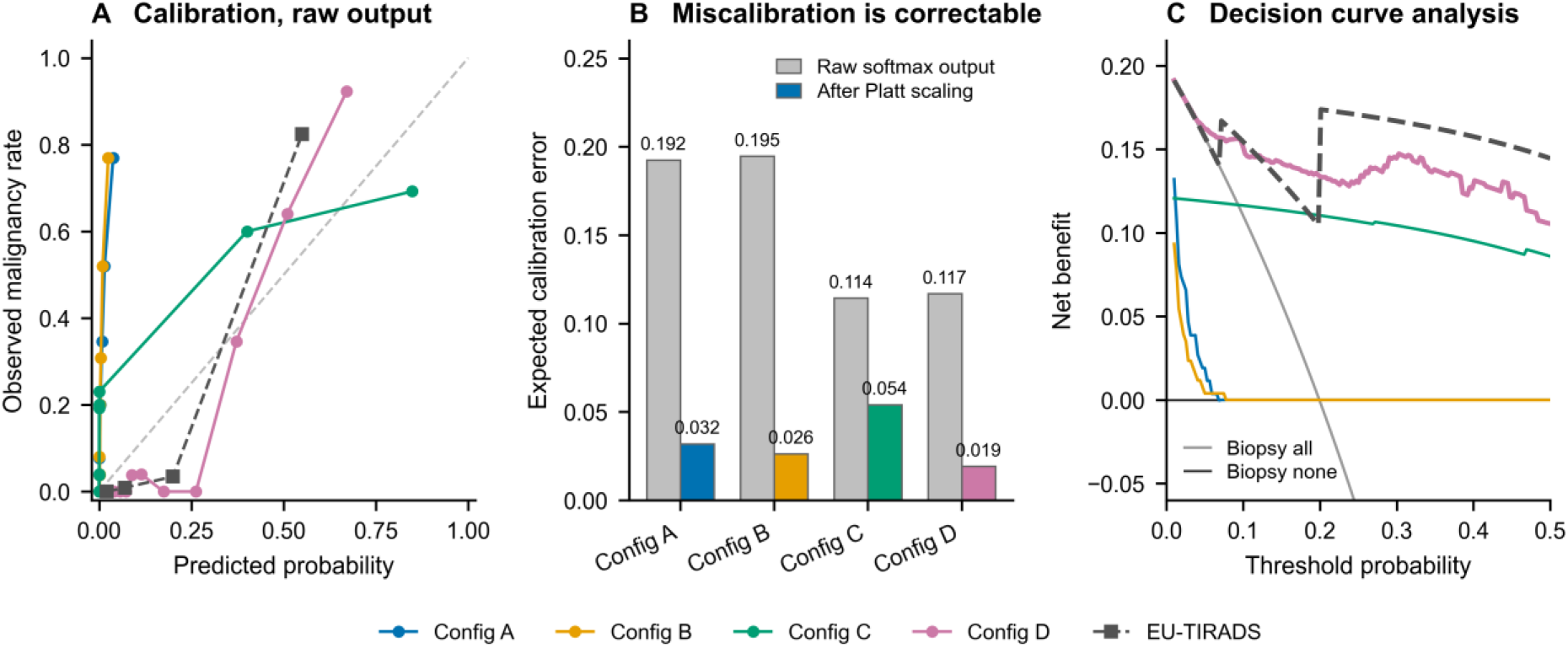
Calibration and clinical utility on the sealed test set. **(A)** Reliability diagrams by decile of predicted probability (ten equal-count bins) for the four configurations, and by category for probabilized EU-TIRADS; the dashed diagonal is perfect calibration. Configs A and B produce probabilities compressed near zero, so their curves rise almost vertically (systematic underestimation of risk). **(B)** Expected calibration error before and after Platt scaling estimated by 5-fold stratified cross-validation within the test set with out-of-fold predictions (seed 42). The error decreases for every configuration (Config D 0.117 → 0.019 at the prespecified seed, 0.019–0.040 across ten seeds) while AUROC changes by no more than 0.006. **(C)** Decision curves over the 0–50% threshold range, with the two reference strategies “biopsy all” and “biopsy none”. Configs A and B return a net benefit of exactly zero above a 10% threshold because no nodule reaches that probability. The stepped appearance of the EU-TIRADS curve reflects the four discrete values the probabilized score can take.

After Platt recalibration estimated by cross-validation within the test set, ECE fell from 0.117 to 0.019 for Config D at the prespecified seed, the low end of a range of 0.019–0.040 across ten seeds (median 0.031); from ≈0.19 to ≈0.03 for Configs A and B; and from 0.114 to 0.054 for Config C. Observed-to-expected ratios returned to ≈1.00, and AUROC changed by ≤ 0.006 (Figure 5B; Supplementary Table S9).

### Clinical utility

Across the clinically relevant threshold range, Config D delivered the highest net benefit at 5% (+0.162) and 15% (+0.142), and probabilized EU-TIRADS at 10% (+0.155) and from 25% upward. At 20%, the threshold coincided with the probability mapped to EU-TIRADS 4 (0.20): with these nodules counted positive, as throughout, Config D led (+0.134 vs +0.104); counted negative, EU-TIRADS would have led (+0.174). Configs A and B, whose probabilities were compressed into [0, 0.08], classified no nodule as positive above a 10% threshold and therefore returned a net benefit of zero (Figure 5C, Table 8).

**Table 8.**
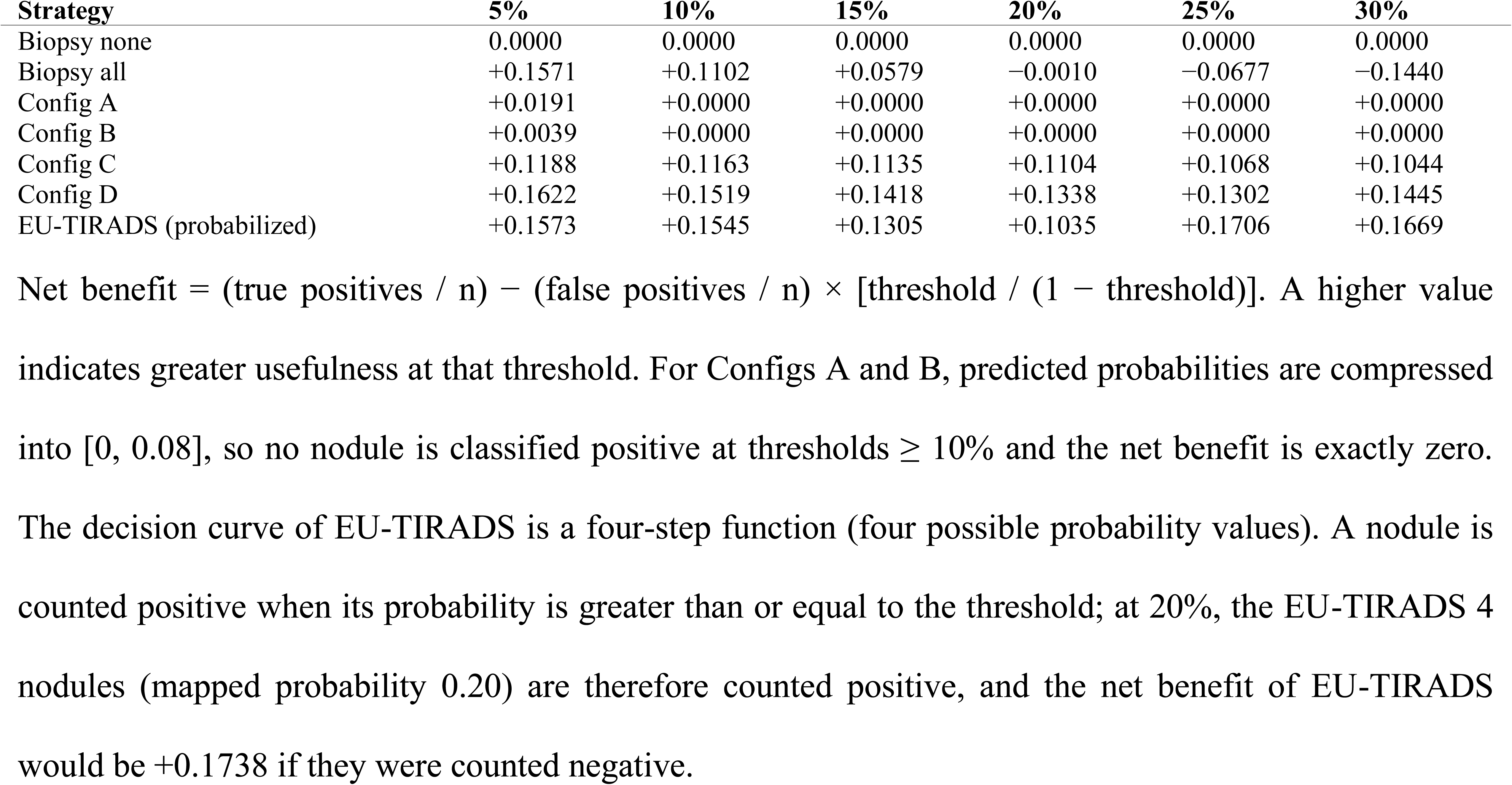
Net benefit by decision curve analysis (sealed test set, n = 256, prevalence 19.9%)

| Strategy | 5% | 10% | 15% | 20% | 25% | 30% |
| --- | --- | --- | --- | --- | --- | --- |
| Biopsy none | 0.0000 | 0.0000 | 0.0000 | 0.0000 | 0.0000 | 0.0000 |
| Biopsy all | +0.1571 | +0.1102 | +0.0579 | -0.0010 | -0.0677 | -0.1440 |
| Config A | +0.0191 | +0.0000 | +0.0000 | +0.0000 | +0.0000 | +0.0000 |
| Config B | +0.0039 | +0.0000 | +0.0000 | +0.0000 | +0.0000 | +0.0000 |
| Config C | +0.1188 | +0.1163 | +0.1135 | +0.1104 | +0.1068 | +0.1044 |
| Config D | +0.1622 | +0.1519 | +0.1418 | +0.1338 | +0.1302 | +0.1445 |
| EU-TIRADS (probabilized) | +0.1573 | +0.1545 | +0.1305 | +0.1035 | +0.1706 | +0.1669 |

Net benefit = (true positives / n) - (false positives / n) × [threshold / (1 - threshold)]. A higher value indicates greater usefulness at that threshold. For Configs A and B, predicted probabilities are compressed into [0, 0.08], so no nodule is classified positive at thresholds $\geq 10\%$ and the net benefit is exactly zero. The decision curve of EU-TIRADS is a four-step function (four possible probability values). A nodule is counted positive when its probability is greater than or equal to the threshold; at 20%, the EU-TIRADS 4 nodules (mapped probability 0.20) are therefore counted positive, and the net benefit of EU-TIRADS would be +0.1738 if they were counted negative.

### Subgroups and robustness

Config D’s AUROC was 0.953 in women and 0.935 in men, 0.948 below and 0.959 from 55 years of age, and 0.949, 0.963 and 0.994 in the Tunis, Mahdia and public-dataset subsets; it increased with nodule size (0.647 below 10 mm, 0.892 at 10–20 mm, 0.992 above 20 mm) (Figure 6); the corresponding estimates for the other three configurations are given in Supplementary Table S10. The microcarcinoma stratum (n = 24) was too small for a reliable estimate and no conclusion is drawn from it. Within-category AUROCs (EU-TIRADS 3, 4 and 5 analyzed separately) were low and imprecise, each stratum containing few events.

**Figure 6.**
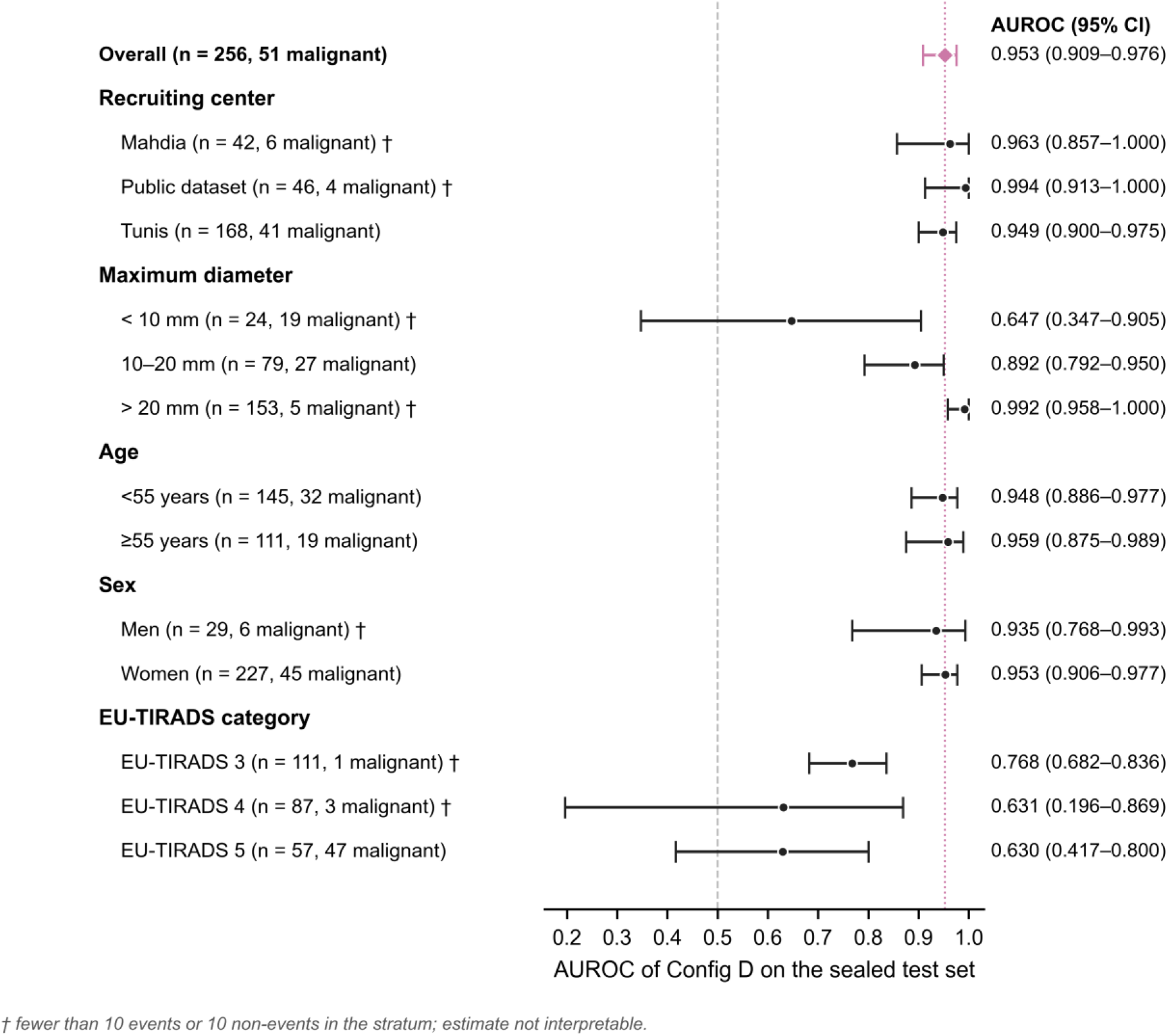
Diagnostic performance of Config D across prespecified subgroups (sealed test set). AUROC with 95% bias-corrected and accelerated bootstrap intervals stratified on outcome (B = 2,000, seed 42). The vertical dotted line marks the overall estimate (0.953) and the dashed line marks chance (0.50). AUROC varies little across sex, age and recruitment source and increases with nodule size. Strata marked † contain fewer than 10 events or fewer than 10 non-events and their estimates are not interpretable; this applies in particular to the within-EU-TIRADS-category analyses.

Verification-bias reweighting by the Begg–Greenes method (overall verification rate 93.3%, 1,456/1,560, the 20 public-dataset images excluded for image reasons lying outside the verification population) changed the EU-TIRADS estimates at ≥ 4 from 97.4% to 97.6% for sensitivity and from 50.5% to 48.2% for specificity (Supplementary Table S11). An algorithmic-fairness audit using the disparate impact ratio (bilateral extension of the four-fifths rule, acceptable range 0.80–1.25) flagged under-classification at the smallest Tunisian center for every configuration (0.45, 0.59, 0.57 and 0.74 for Configs A–D) and, for Config D, on the public-dataset subset (0.68) and in patients aged 60 years and over (0.51–0.54) (Supplementary Table S12). Reinstating the 104 indeterminate Tunisian nodules moved the cohort-level ordinal AUROC of EU-TIRADS from 0.921 to 0.912 if all were assumed benign and to 0.867 if all were assumed malignant (Supplementary Table S13).

### EU-TIRADS-anchored decision guide

Applied to the sealed test set, the decision guide (Figure 7) left the 112 EU-TIRADS 2–3 nodules (1 cancer) and the 57 EU-TIRADS 5 nodules (47 cancers) on their usual pathway; Config D called none of the category 2–3 nodules malignant, so the safety net was never triggered. None of the 87 EU-TIRADS 4 nodules (3 cancers) carried a high-risk descriptor, as the definition of the category implies. At the threshold fixed on validation data, FNA would have been deferred for 71 of the 84 benign category 4 nodules and for 2 of the 3 cancers; at the Youden threshold of the test set, for 68 of the 84 benign nodules and 1 of the 3 cancers.

**Figure 7.**
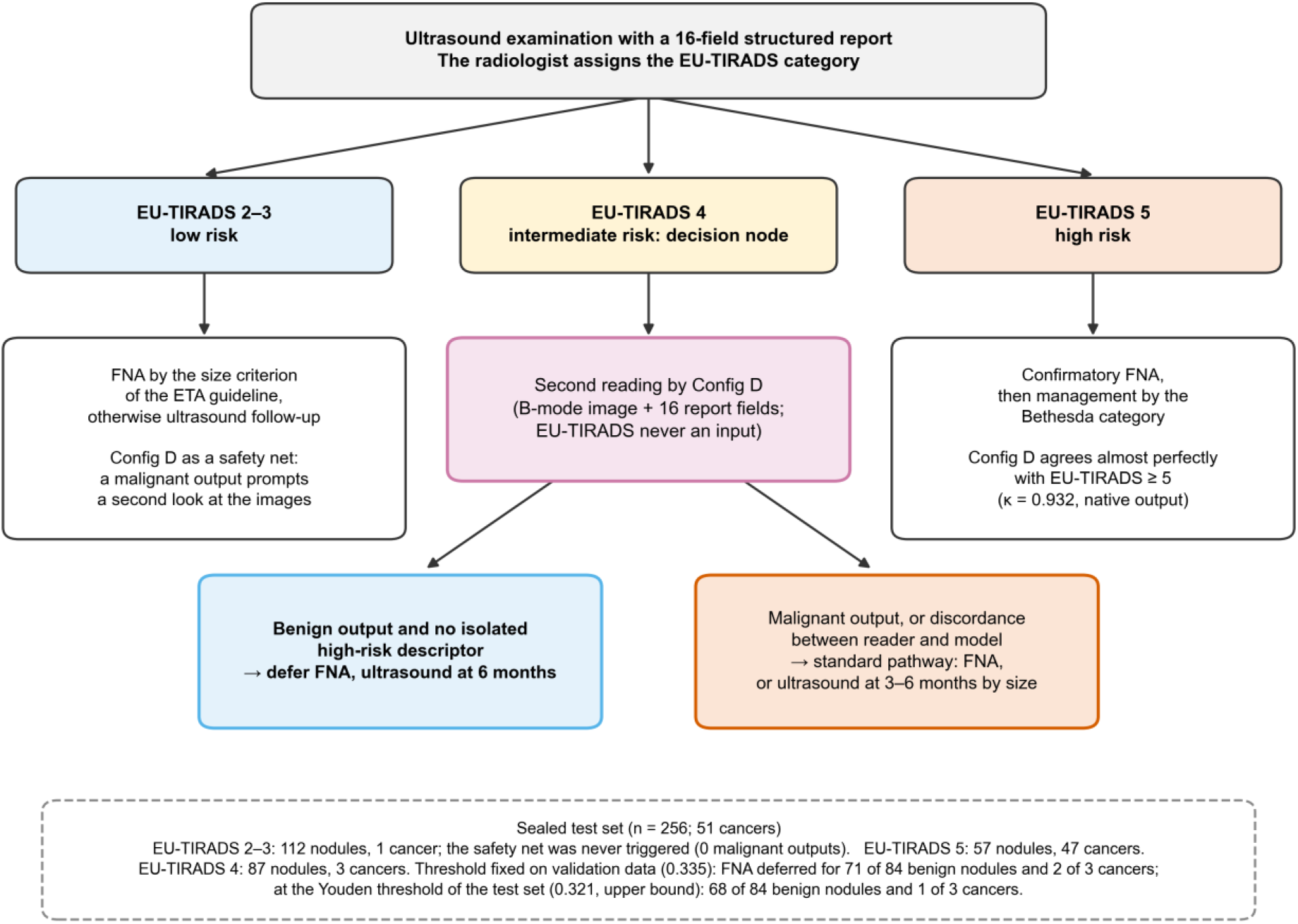
Proposed EU-TIRADS-anchored decision guide for Config D as a second reader, applied to the sealed test set (n = 256; 51 cancers). EU-TIRADS, assigned by the radiologist, remains the first-line classifier. In categories 2–3, management follows the size criteria of the European Thyroid Association guideline and Config D acts only as a safety net; in category 5, confirmatory FNA is kept, Config D agreeing almost perfectly with the EU-TIRADS ≥ 5 dichotomy (κ = 0.932 at its native threshold). In category 4, a benign output with no isolated high-risk descriptor (marked hypoechogenicity, angular or lobulated margins, microcalcifications, taller-than-wide shape) defers FNA in favor of ultrasound at 6 months, whereas a malignant output or a discordance between reader and model keeps the standard pathway. The counts at the bottom come from the sealed test set: at the threshold fixed on validation data (0.335), FNA would have been deferred for 71 of the 84 benign category 4 nodules and for 2 of its 3 cancers; at the Youden threshold of the test set (0.321, an upper bound), for 68 of 84 and 1 of 3. The guide is exploratory and has not been validated prospectively. ETA = European Thyroid Association, FNA = fine-needle aspiration.

## Discussion

In a Tunisian multicenter cohort of 1,456 nodules with a test set of 256 nodules sealed before training, a LoRA-fine-tuned medical vision–language model reached an AUROC of 0.953 when given the ultrasound image together with the radiologist’s structured report. This was statistically indistinguishable from expert-rated EU-TIRADS (0.948) and within the prespecified non-inferiority margin. Configurations limited to the image and quantitative biometry, without qualitative descriptors, scored significantly below the human score, so it was the combination of the image with the structured report that reached expert level. The result of clinical interest is therefore not a gain in discrimination but a different operating point. Near maximal sensitivity, the model converted the excess sensitivity of EU-TIRADS into roughly 34 percentage points of specificity at the test-derived threshold, avoiding 73% of the false positives of the score for one additional cancer not immediately aspirated, and into 35 points (72 of 94 false positives, two cancers) at the threshold fixed on validation data. This gain is defined against the ≥ 4 threshold. Against the ≥ 5 threshold, Config D did not classify more nodules correctly: its native output reproduced that dichotomy almost exactly (κ = 0.932), and at the Youden and validation-derived thresholds it detected one or two additional cancers at the cost of 12 to 15 additional false positives. Of the 69 false positives reclassified at the Youden threshold, 68 were in category 4, so the specific contribution of the model lies within that category, where the test set held only three cancers and cannot establish a safe miss rate. We kept ≥ 4 as the comparator because it is the risk-classifier dichotomy of EU-TIRADS; the size-adapted indication for FNA is not a malignancy classifier in an aspirated cohort and would have missed 27 of the 51 cancers, most of them small. Finally, the binary NRI weights a missed cancer (1/51) and a spared benign nodule (1/205) by the size of their classes rather than by their clinical cost; net benefit, which states that exchange rate explicitly, is the more relevant summary.

### Discrimination relative to EU-TIRADS

EU-TIRADS is itself a structured expert reading of the image, derived from the same qualitative descriptors that Config D receives, so discrimination close to that of the score is the expected result for a model given that report. Only one image per nodule was available, which limits the diversity of visual features the model could use. The precision achieved on the paired difference (standard error 0.010 on 256 nodules with 51 events) still admits a deficit of up to 0.016 in favor of EU-TIRADS, and roughly 1,000 nodules would be needed to halve that interval [52], so the absence of a significant difference cannot be read as formal equivalence. Lastly, excluding the global EU-TIRADS category from the model inputs, a precaution rarely made explicit [26], removes input circularity but forces the model to reconstruct that information from the descriptors it receives, which may itself blunt any apparent advantage.

### What Config D is

Config D consumes a 16-field structured input, eleven fields of which are the qualitative sonographic descriptors from which EU-TIRADS is derived. It is therefore not an autonomous image reader but a decision-support layer that standardizes the step from a structured report to a risk estimate, which is the step where interobserver variability is documented [6,7]. Its non-inferiority to EU-TIRADS is accordingly a statement about the reproducibility of scoring, not about machine vision replacing radiologists. Config A and Config C differed by 0.009 in AUROC; they differ by the removal of the auxiliary EU-TIRADS target but also by the corrected logit extraction and the chain-of-thought supervision. Co-supervision therefore inflates performance only to a clinically negligible degree, but this bound applies to the configurations without qualitative descriptors and cannot be transferred directly to Config D.

### Comparison with the literature

ThyNet reported an AUROC of 0.92 in a multicenter Chinese cohort [14], and the multimodal ThyGPT reported 0.95 with a reduction in FNA of more than 40% across nine hospitals [15]. A meta-analysis of 27 studies with external datasets found a summary AUROC of 0.919 (0.900–0.932), with pooled sensitivity of 0.87 and specificity of 0.83, and called for evaluation outside Asian cohorts [12]. Our point estimate lies above that pooled bound, but the comparison is illustrative rather than inferential: it comes from a single cohort of limited size, whereas the pooled figure aggregates heterogeneous populations and systems, several of them validated externally. External validation, not yet available, is the benchmark against which the internal estimate of 0.953 will have to be judged. Conversely, the AUROC of EU-TIRADS in our cohort (0.948) is higher than the 0.85 reported in a single-center series of 4,238 aspirated nodules (8.9% malignant) [53] and than pooled estimates across risk stratification systems [54]. This reflects enrichment in aspirated and operated nodules and the exclusion of indeterminate cytology, a spectrum effect quantified below.

### Calibration

Raw calibration was imperfect for every configuration, but correctable. All four rejected perfect calibration, and two produced probabilities less informative than the prevalence itself. This is a well-described property of softmax outputs that have not been recalibrated [47], and in our data it was largely reversible: Platt scaling reduced the calibration error of Config D three-to sixfold, depending on the fold draw, without affecting discrimination (Supplementary Table S9). The calibration of EU-TIRADS was mixed, with a Brier score slightly lower than that of Config D (0.075 vs 0.080) but a higher expected calibration error (0.144 vs 0.117) and a steeper slope (2.78 vs 2.51). These comparisons should be read with caution, because the probabilities assigned to EU-TIRADS are working values chosen by the authors whereas the model outputs were raw softmax values. Discrimination and utility can also diverge: Configs A and B, with AUROCs close to 0.89, returned a net benefit of zero above a 10% threshold because their probabilities were compressed, and the lead of EU-TIRADS at isolated thresholds of the decision curve reflects the steps of a four-level score, including a tie at exactly 20%, rather than a general superiority. A deployable system would have to be recalibrated on an independent validation set, and a configuration should be chosen for the quality of its calibration as much as for its discrimination [49].

### Strengths

The evaluation rests on a test set sealed and checksummed before model training, with partitioning at patient level; a strictly paired expert comparator from the same population; explicit exclusion of the comparator score from model inputs; prespecified hypotheses, thresholds and non-inferiority margin; a prespecified sensitivity analysis (Config C) bounding co-supervision bias; and the joint evaluation of discrimination, calibration and clinical utility recommended for prediction models [49], rarely reported together in this field. To our knowledge, this is the first evaluation of a multimodal medical foundation model for thyroid nodule risk stratification in a Tunisian, and more broadly North African, cohort. With ≈3.5% of parameters trained and inference in bfloat16 on a single graphics processing unit, the approach can run on modest local hardware.

### Limitations

The EU-TIRADS category, although never an input, was an auxiliary learning target in Configs A, B and D, and the auxiliary confidence target of these configurations was set, for benign nodules, from EU-TIRADS category ranges; Config C, which omits this target, bounds its effect on performance. The reference standard was mainly cytological: 66 Tunisian nodules classified malignant on Bethesda V were not systematically confirmed by histology, and given the 65–75% malignancy risk of that category, a fraction may be benign, a misclassification that affects model and comparator alike. In the Tunisian centers, histology was available for 11 nodules, all malignant, so the ten Bethesda III–IV nodules included are all malignant. Excluding the other indeterminate nodules evaluates both tests on a distribution enriched in clear-cut cases: reinstating the 104 indeterminate Tunisian nodules moves the cohort-level ordinal AUROC of EU-TIRADS from 0.921 to 0.912 if all are assumed benign, or to 0.867 if all are assumed malignant (Supplementary Table S13), and the Begg–Greenes reweighting relies on a missing-at-random assumption that may not hold when verification depends on the surgical decision. The four configurations were evaluated on the same test set; the comparisons with EU-TIRADS were adjusted for multiplicity, and the advantage of Config D over the other configurations should be confirmed in an independent cohort. The respective contributions of the image and of the structured descriptors were not separated by a modality ablation of the fine-tuned model. The cohort is imbalanced across sources (Tunis 84.7% of the cohort and 65.6% of the test set), partly because the first 750 records, all from Tunis in 2019–2022, were assigned to training, which also introduces a temporal and spectrum difference between partitions; this is not equivalent to external validation by center, and the disparate impact ratio was out of range at the smallest center. The 103 public-dataset images come from a different scanner fleet and population, with a histological reference standard and EU-TIRADS ratings assigned on stored images selected for their sharpness and quality, which qualifies the Tunisian character of the cohort; this selection may have made the task easier on that subset (Config D AUROC 0.994). The probabilities assigned to the EU-TIRADS categories are conventional, so the Brier score, calibration and net benefit of EU-TIRADS depend on that convention, whereas its AUROC does not. Predicted probabilities were kept only for the test set, so recalibration had to be estimated by cross-validation within it, and threshold-free validation metrics could not be computed. Each nodule was rated once, by the radiologist of its center, so the interobserver variability of EU-TIRADS and of the descriptors was not measured, and the model analyzed single 2D B-mode images without elastography or three-dimensional acquisition. Finally, the predictive values (positive 78.9%, negative 97.0% at the native threshold) depend on a cohort prevalence of 18.7% driven by aspiration-based recruitment and do not transfer to unselected nodule populations, and no prospective external validation has been performed [25]. Confidence intervals were computed at nodule level and do not account for the clustering of nodules within the minority of patients who contributed several nodules.

### Clinical implications and proposed decision guide

The results support use as a second reader after the ultrasound examination, not as autonomous image interpretation, and the EU-TIRADS-anchored decision guide of Figure 7 makes this role explicit. EU-TIRADS remains the first-line classifier assigned by the radiologist. In categories 2–3, management follows the size criteria of the guidelines and the model serves only as a safety net that prompts a second look at the images; it was never triggered on the test set, but it did not flag the only cancer in these categories either. In category 5, the native output of the model agreed almost perfectly with the EU-TIRADS ≥ 5 dichotomy (κ = 0.932), so it would confirm rather than modify the indication. Category 4, the gray zone where specificity is lowest, is the decision node: a benign output with no isolated high-risk descriptor could support deferring aspiration in favor of ultrasound surveillance at six months. Restricted to category 4, this rule would have deferred 71 of the 84 unnecessary aspirations at the threshold derived on the validation partition alone (0.335), which never saw the test data, for two of the three cancers in the category, and 68 of 84 for one of three at the Youden threshold, against a comparator that already missed one cancer in 51. With three cancers in the category, these counts are descriptive and do not estimate a miss rate. The lower rate of positive calls in patients aged 60 years and over (disparate impact ratio 0.51–0.54) partly reflects the younger age of patients with cancer in this cohort (median 46 vs 53 years) but would have to be monitored in any deployment. Any implementation presupposes a 16-field structured report, refusal to infer when a descriptor is missing, output of a probability with its confidence interval rather than a binary label, and local prospective revalidation. The decision threshold is itself an adjustable setting rather than a fixed property of the model. On the test set, lowering it to 0.10 recovered the sensitivity of EU-TIRADS ≥ 4 but at the price of more false positives than the score, whereas raising it to 0.40 or more spared further aspirations only at the cost of a steep rise in missed cancers (Supplementary Table S8); a center can therefore set it according to the balance it accepts between missed cancers and aspirations. Because the LoRA adapters are light and inexpensive to train, the model could likewise be refined on the data of a single center, with its own radiologists, reporting habits and scanners; such local adaptation would probably fit that setting better than one global model, provided it is confirmed by local validation. Such a system would fall within the high-risk category of Regulation (EU) 2024/1689 [55] and would require compliance with data-protection law and TRIPOD+AI reporting [29].

### Future directions

The next steps are, in order: formal recalibration on an independent validation set; an ablation quantifying the respective contributions of the image and of the tabular input; targeted evaluation in Bethesda III–IV nodules, the population in which a probabilistic tool available before aspiration would be most useful and which was excluded here; and a prospective, preregistered multicenter validation of at least 2,000 consecutive nodules across five or more centers with systematic histological confirmation. Richer acoustic information (elastography, Doppler, multi-image protocols) is a realistic medium-term avenue. Because only a minority of the 26 units approached had an exploitable picture archiving and communication system, federated learning combined with differential privacy offers a practical route to multicenter enrichment without centralizing data.

## Conclusion

In a Tunisian multicenter cohort with a sealed test set, a parameter-efficiently fine-tuned medical vision– language model matched, but did not surpass, the discrimination of expert-rated EU-TIRADS. It converted the excess sensitivity of the score into a substantial gain in specificity, reclassifying as negative about three of every four EU-TIRADS false positives (72 of 94 at a threshold fixed on validation data, for two cancers newly missed), an estimate based on only three cancers in category 4, and it agreed closely with the ≥ 5 threshold. Its performance rested on the combination of the image with the radiologist’s structured descriptors, and its raw probabilities, initially miscalibrated, were largely corrected by Platt scaling. These findings support further development as a second-reader, report-standardizing decision aid; prospective external validation and formal recalibration are prerequisites for clinical use.

## Declarations

### Ethics approval

Approved by the Medical Ethics Committee of Taher Sfar University Hospital, Mahdia, Tunisia (ref. CEM-2025-10-04). Retrospective study on pseudonymized data; individual informed consent was waived under Tunisian decree no. 2007-2609. Conducted in accordance with the Declaration of Helsinki. The study was not registered and no protocol was published; hypotheses, thresholds, the non-inferiority margin and subgroups were fixed before the test set was analyzed. Patients and the public were not involved in the design, conduct or reporting of this study.

### Consent for publication

Not applicable.

### Funding

This study received no institutional, public or industrial funding. Computing and development costs were borne personally by M.N.B.A. and H.M.

### Competing interests

The authors declare no competing interests. No author has a financial relationship with Google, Google DeepMind or any provider of the models used.

### Data availability

The coded clinical variables of the 1,580 documented nodules, the locked per-nodule prediction files of the four configurations, the sealed-split definition and its SHA-256 checksum, the fine-tuning configuration files and the training log are available from the corresponding author on reasonable request, subject to the approvals of the participating centers and to Tunisian personal-data legislation. The statistical analysis code, which regenerates every reported value from these files, is available with the data. Patient ultrasound images from the Tunisian centers cannot be shared, and the fine-tuned LoRA adapter weights are not released. The international public component of the training data is openly available from Hou et al. (figshare, DOI 10.6084/m9.figshare.27021604.v1; CC BY 4.0) [34].

### Model and code availability

The fine-tuned LoRA adapter weights and the training and inference code are not released. The base model, MedGemma 1.5 4B (google/medgemma-1.5-4b-it), is publicly available from its developers under their own license [19]. All hyperparameters, prompt structure, preprocessing steps and analysis procedures required to reimplement the study are specified in the Materials and Methods and the Supplementary Appendix.

### Author contributions

M.N.B.A.: conceptualization, data curation, formal analysis, investigation, methodology, visualization, writing — original draft, review and editing. N.L.: conceptualization, supervision, clinical methodology, validation, writing — review and editing. H.M.: software, model training infrastructure, methodology, validation, review and editing. E.M.: investigation (ultrasound acquisition and EU-TIRADS rating, Tunis) and resources. S.H.: investigation (cytopathology and histopathology, Tunis) and resources. K.B.R.: investigation (ultrasound acquisition and EU-TIRADS rating, Mahdia) and resources. M.K.E.: investigation (cytopathology, Sousse, paired with the Mahdia imaging center) and resources. M.H.: review and editing. All authors read and approved the final manuscript and agree to be accountable for all aspects of the work.

## Acknowledgments

The authors thank the radiology and pathology teams of the participating centers, and Dr Mahmoud Ben Khlifa and Dr Moemen Abdallah for their contribution to the project. They gratefully acknowledge Hou and colleagues for making their annotated thyroid ultrasound dataset openly available under CC BY 4.0 [34], which contributed part of the training data, and Google DeepMind and Google Research for releasing the MedGemma model family under open weights [18,19], without which this work would not have been feasible on local hospital infrastructure. Neither organization had any role in the design, conduct, analysis, interpretation or reporting of this study.

## Thesis

This work is part of the doctoral thesis in medicine of M.N.B.A. (Faculty of Medicine of Monastir, University of Monastir, 2026), which has not been published in a journal.

## Use of AI tools

Large language model assistance was used for language editing and for assistance with code writing and document formatting. All analyses were executed by the authors from source data; all results, references and interpretations were verified by the authors, who take full responsibility for the content.

## Notes

### Competing Interest Statement

The authors have declared no competing interest.

### Author Declarations

Medical Ethics Committee of Taher Sfar University Hospital, Mahdia, Tunisia gave ethical approval for this work.

## References

1. Dean DS, Gharib H. Epidemiology of thyroid nodules. Best Pract Res Clin Endocrinol Metab. 2008;22(6):901–11. doi:10.1016/j.beem.2008.09.019

2. Durante C, Hegedüs L, Czarniecka A, Paschke R, Russ G, Schmitt F, et al. 2023 European Thyroid Association clinical practice guidelines for thyroid nodule management. Eur Thyroid J. 2023;12(5):e230067. doi:10.1530/ETJ-23-0067

3. Russ G, Bonnema SJ, Erdogan MF, Durante C, Ngu R, Leenhardt L. European Thyroid Association guidelines for ultrasound malignancy risk stratification of thyroid nodules in adults: the EU-TIRADS. Eur Thyroid J. 2017;6(5):225–37. doi:10.1159/000478927

4. Kim PH, Suh CH, Baek JH, Chung SR, Choi YJ, Lee JH. Unnecessary thyroid nodule biopsy rates under four ultrasound risk stratification systems: a systematic review and meta-analysis. Eur Radiol. 2021;31(5):2877–85. doi:10.1007/s00330-020-07384-6

5. Castellana M, Castellana C, Treglia G, Giorgino F, Giovanella L, Russ G, et al. Performance of five ultrasound risk stratification systems in selecting thyroid nodules for FNA. J Clin Endocrinol Metab. 2020;105(5):1659–69. doi:10.1210/clinem/dgz170

6. Alyami J, Almutairi FF, Aldoassary S, Albeshry A, Almontashri A, Abounassif M, et al. Interobserver variability in ultrasound assessment of thyroid nodules. Medicine (Baltimore). 2022;101(41):e31106. doi:10.1097/MD.0000000000031106

7. Grani G, Lamartina L, Cantisani V, Maranghi M, Lucia P, Durante C. Interobserver agreement of various thyroid imaging reporting and data systems. Endocr Connect. 2018;7(1):1–7. doi:10.1530/EC-17-0336

8. Vaccarella S, Franceschi S, Bray F, Wild CP, Plummer M, Dal Maso L. Worldwide thyroid-cancer epidemic? The increasing impact of overdiagnosis. N Engl J Med. 2016;375(7):614–7. doi:10.1056/NEJMp1604412

9. Bray F, Laversanne M, Sung H, Ferlay J, Siegel RL, Soerjomataram I, et al. Global cancer statistics 2022: GLOBOCAN estimates of incidence and mortality worldwide for 36 cancers in 185 countries. CA Cancer J Clin. 2024;74(3):229–63. doi:10.3322/caac.21834

10. Khiari H, Arfaoui E, Mahjoub N, Henchiri S, Sliti A, Hsairi M. Estimated cancer incidence in Northern Tunisia in 2023: Northern Tunisia Cancer Registry. Asian Pac J Cancer Prev. 2024;25(12):4359–69. doi:10.31557/APJCP.2024.25.12.4359

11. Nejadghaderi SA, Moghaddam SS, Azadnajafabad S, Rezaei N, Rezaei N, Tavangar SM, et al. Burden of thyroid cancer in North Africa and Middle East 1990–2019. Front Oncol. 2022;12:955358. doi:10.3389/fonc.2022.955358. Erratum in: Front Oncol. 2023;13:1208646

12. Gatta E, Gatta R, Morandi R, Isoli S, Corvaglia S, Vetrugno S, et al. Machine learning for diagnosis of malignant thyroid nodules based on thyroid ultrasound: systematic review and meta-analysis of studies with external datasets. Eur J Radiol Open. 2026;16:100716. doi:10.1016/j.ejro.2025.100716

13. Zhan J, Zhang J, Zhu S, Ni L, Zhang C, Hu J. Diagnostic performance of ultrasound characteristics-based artificial intelligence models for thyroid nodules: a systematic review and meta-analysis. Front Oncol. 2025;15:1614603. doi:10.3389/fonc.2025.1614603

14. Peng S, Liu Y, Lv W, Liu L, Zhou Q, Yang H, et al. Deep learning-based artificial intelligence model to assist thyroid nodule diagnosis and management: a multicentre diagnostic study. Lancet Digit Health. 2021;3(4):e250–9. doi:10.1016/S2589-7500(21)00041-8

15. Yao J, Wang Y, Lei Z, Wang K, Feng N, Dong F, et al. Multimodal GPT model for assisting thyroid nodule diagnosis and management. NPJ Digit Med. 2025;8(1):245. doi:10.1038/s41746-025-01652-9

16. Zhang S, Xu Y, Usuyama N, Xu H, Bagga J, Tinn R, et al. A multimodal biomedical foundation model trained from fifteen million image–text pairs. NEJM AI. 2025;2(1):AIoa2400640. doi:10.1056/AIoa2400640

17. Li C, Wong C, Zhang S, Usuyama N, Liu H, Yang J, et al. LLaVA-Med: training a large language-and-vision assistant for biomedicine in one day. Adv Neural Inf Process Syst. 2023;36. arXiv:2306.00890. doi:10.48550/arXiv.2306.00890

18. Sellergren A, Kazemzadeh S, Jaroensri T, Kiraly A, Traverse M, Kohlberger T, et al. MedGemma technical report. arXiv:2507.05201 [Preprint]. 2025. doi:10.48550/arXiv.2507.05201

19. Sellergren A, Gao C, Mahvar F, Kohlberger T, Jamil F, Traverse M, et al. MedGemma 1.5 technical report. arXiv:2604.05081 [Preprint]. 2026. doi:10.48550/arXiv.2604.05081

20. Hu EJ, Shen Y, Wallis P, Allen-Zhu Z, Li Y, Wang S, et al. LoRA: low-rank adaptation of large language models. In: Proceedings of the 10th International Conference on Learning Representations (ICLR); 2022. arXiv:2106.09685. doi:10.48550/arXiv.2106.09685

21. Dutt R, Ericsson L, Sanchez P, Tsaftaris SA, Hospedales T. Parameter-efficient fine-tuning for medical image analysis: the missed opportunity. Proc Mach Learn Res. 2024;250:406–25. arXiv:2305.08252. doi:10.48550/arXiv.2305.08252

22. Sng GGR, Xiang Y, Lim DYZ, Tung JYM, Tan JH, Chng CL. A multimodal large language model as an end-to-end classifier of thyroid nodule malignancy risk: usability study. JMIR Form Res. 2025;9:e70863. doi:10.2196/70863

23. Li YH, Cheng YC, Chang CY, Lee IT. Optimizing a multimodal large language model for ultrasound-based thyroid nodule malignancy classification: a comparative study of few-shot learning, prompt engineering, and fine-tuning. Diagnostics (Basel). 2026;16(12):1931. doi:10.3390/diagnostics16121931

24. Andaur Navarro CL, Damen JAA, Takada T, Nijman SWJ, Dhiman P, Ma J, et al. Completeness of reporting of clinical prediction models developed using supervised machine learning: a systematic review. BMC Med Res Methodol. 2022;22(1):12. doi:10.1186/s12874-021-01469-6

25. Nagendran M, Chen Y, Lovejoy CA, Gordon AC, Komorowski M, Harvey H, et al. Artificial intelligence versus clinicians: systematic review of design, reporting standards, and claims of deep learning studies. BMJ. 2020;368:m689. doi:10.1136/bmj.m689

26. Kapoor S, Narayanan A. Leakage and the reproducibility crisis in machine-learning-based science. Patterns (N Y). 2023;4(9):100804. doi:10.1016/j.patter.2023.100804

27. Bossuyt PM, Reitsma JB, Bruns DE, Gatsonis CA, Glasziou PP, Irwig L, et al. STARD 2015: an updated list of essential items for reporting diagnostic accuracy studies. BMJ. 2015;351:h5527. doi:10.1136/bmj.h5527

28. Cohen JF, Korevaar DA, Altman DG, Bruns DE, Gatsonis CA, Hooft L, et al. STARD 2015 guidelines for reporting diagnostic accuracy studies: explanation and elaboration. BMJ Open. 2016;6(11):e012799. doi:10.1136/bmjopen-2016-012799

29. Collins GS, Moons KGM, Dhiman P, Riley RD, Beam AL, Van Calster B, et al. TRIPOD+AI statement: updated guidance for reporting clinical prediction models that use regression or machine learning methods. BMJ. 2024;385:e078378. doi:10.1136/bmj-2023-078378

30. Gallifant J, Afshar M, Ameen S, Aphinyanaphongs Y, Chen S, Cacciamani G, et al. The TRIPOD-LLM reporting guideline for studies using large language models. Nat Med. 2025;31(1):60–9. doi:10.1038/s41591-024-03425-5

31. Mongan J, Moy L, Kahn CE Jr. Checklist for Artificial Intelligence in Medical Imaging (CLAIM): a guide for authors and reviewers. Radiol Artif Intell. 2020;2(2):e200029. doi:10.1148/ryai.2020200029

32. Tejani AS, Klontzas ME, Gatti AA, Mongan JT, Moy L, Park SH, et al. Checklist for Artificial Intelligence in Medical Imaging (CLAIM): 2024 update. Radiol Artif Intell. 2024;6(4):e240300. doi:10.1148/ryai.240300

33. World Medical Association. World Medical Association Declaration of Helsinki: ethical principles for medical research involving human participants. JAMA. 2025;333(1):71–4. doi:10.1001/jama.2024.21972

34. Hou X, Hua M, Zhang W, Ji J, Zhang X, Jiang H, et al. An ultrasonography of thyroid nodules dataset with pathological diagnosis annotation for deep learning. Sci Data. 2024;11(1):1272. doi:10.1038/s41597-024-04156-5

35. Usher-Smith JA, Sharp SJ, Griffin SJ. The spectrum effect in tests for risk prediction, screening, and diagnosis. BMJ. 2016;353:i3139. doi:10.1136/bmj.i3139

36. Ali SZ, Baloch ZW, Cochand-Priollet B, Schmitt FC, Vielh P, VanderLaan PA. The 2023 Bethesda System for Reporting Thyroid Cytopathology. Thyroid. 2023;33(9):1039–44. doi:10.1089/thy.2023.0141

37. Begg CB, Greenes RA. Assessment of diagnostic tests when disease verification is subject to selection bias. Biometrics. 1983;39(1):207–15. doi:10.2307/2530820

38. Buderer NMF. Statistical methodology: I. Incorporating the prevalence of disease into the sample size calculation for sensitivity and specificity. Acad Emerg Med. 1996;3(9):895–900. doi:10.1111/j.1553-2712.1996.tb03538.x

39. Alseddeeqi E, Baharoon R, Mohamed R, Ghaith J, Al-Helali A, Ahmed LA. Thyroid malignancy among patients with thyroid nodules in the United Arab Emirates: a five-year retrospective tertiary centre analysis. Thyroid Res. 2018;11(1):17. doi:10.1186/s13044-018-0061-x

40. Al-Sharafi BA, AlSanabani JA, Alboany IM, Shamsher AM. Thyroid cancer among patients with thyroid nodules in Yemen: a three-year retrospective study in a tertiary center and a specialty clinic. Thyroid Res. 2020;13(1):8. doi:10.1186/s13044-020-00082-x

41. Obuchowski NA. ROC analysis. AJR Am J Roentgenol. 2005;184(2):364–72. doi:10.2214/ajr.184.2.01840364

42. Wilson EB. Probable inference, the law of succession, and statistical inference. J Am Stat Assoc. 1927;22(158):209–12. doi:10.1080/01621459.1927.10502953

43. Castellana M, Grani G, Radzina M, Guerra V, Giovanella L, Deandrea M, et al. Performance of EU-TIRADS in malignancy risk stratification of thyroid nodules: a meta-analysis. Eur J Endocrinol. 2020;183(3):255–64. doi:10.1530/EJE-20-0204

44. Saito T, Rehmsmeier M. The precision-recall plot is more informative than the ROC plot when evaluating binary classifiers on imbalanced datasets. PLoS One. 2015;10(3):e0118432. doi:10.1371/journal.pone.0118432

45. Efron B. Better bootstrap confidence intervals. J Am Stat Assoc. 1987;82(397):171–85. doi:10.1080/01621459.1987.10478410

46. Spiegelhalter DJ. Probabilistic prediction in patient management and clinical trials. Stat Med. 1986;5(5):421–33. doi:10.1002/sim.4780050506

47. Guo C, Pleiss G, Sun Y, Weinberger KQ. On calibration of modern neural networks. Proc Mach Learn Res. 2017;70:1321–30. arXiv:1706.04599. doi:10.48550/arXiv.1706.04599

48. Vickers AJ, Elkin EB. Decision curve analysis: a novel method for evaluating prediction models. Med Decis Making. 2006;26(6):565–74. doi:10.1177/0272989X06295361

49. Van Calster B, Collins GS, Vickers AJ, Wynants L, Kerr KF, Barreñada L, et al. Evaluation of performance measures in predictive artificial intelligence models to support medical decisions: overview and guidance. Lancet Digit Health. 2025;7(12):100916. doi:10.1016/j.landig.2025.100916

50. DeLong ER, DeLong DM, Clarke-Pearson DL. Comparing the areas under two or more correlated receiver operating characteristic curves: a nonparametric approach. Biometrics. 1988;44(3):837– 45. doi:10.2307/2531595

51. Pencina MJ, D’Agostino RB Sr, D’Agostino RB Jr, Vasan RS. Evaluating the added predictive ability of a new marker: from area under the ROC curve to reclassification and beyond. Stat Med. 2008;27(2):157–72. doi:10.1002/sim.2929

52. Riley RD, Debray TPA, Collins GS, Archer L, Ensor J, van Smeden M, et al. Minimum sample size for external validation of a clinical prediction model with a binary outcome. Stat Med. 2021;40(19):4230–51. doi:10.1002/sim.9025

53. Özdemir M, Türk G, Bilgili M, Akay E, Koç A. Comparison of diagnostic performances of ATA guidelines, ACR-TIRADS, and EU-TIRADS and modified K-TIRADS: a single center study of 4238 thyroid nodules. Exp Clin Endocrinol Diabetes. 2025;133(2):98–104. doi:10.1055/a-2498-7952

54. Kim JS, Kim BG, Stybayeva G, Hwang SH. Diagnostic performance of various ultrasound risk stratification systems for benign and malignant thyroid nodules: a meta-analysis. Cancers (Basel). 2023;15(2):424. doi:10.3390/cancers15020424

55. European Parliament, Council of the European Union. Regulation (EU) 2024/1689 of 13 June 2024 laying down harmonised rules on artificial intelligence (Artificial Intelligence Act). Off J Eur Union. 2024;L 2024/1689. Available from: http://data.europa.eu/eli/reg/2024/1689/oj

