## supplemental File for "Fine-tuned Medical Vision–Language Model (MedGemma 1.5 4B) with and without Structured Radiologist Descriptors versus EU-TIRADS for Malignancy Risk Stratification of Thyroid Nodules at Ultrasound: A Tunisian Multicenter Diagnostic Accuracy Study"

---

#### Contents


- Section A. Reporting checklists
- Supplementary Table S1. STARD 2015 checklist
- Supplementary Table S2. TRIPOD+AI checklist (key items)
- Supplementary Table S3. CLAIM 2024 checklist (key items)
- Section B. Variables and prompt construction
- Supplementary Table S4. Variables collected and their use as model inputs
- Section C. Training protocol
- Supplementary Table S5. Shared training configuration
- Supplementary Table S6. Differences between the four configurations
- Supplementary Table S7. Checkpoint selection and validation performance
- Section D. Methodological definitions
- Section E. Supplementary results
- Supplementary Table S8. Config D across decision thresholds: cancers detected and aspirations indicated (sealed test set)
- Supplementary Table S9. Recalibration by Platt scaling (internal cross-validation on the sealed test set)
- Supplementary Table S10. AUROC by prespecified subgroup, all four configurations (sealed test set)
- Supplementary Table S11. Verification-bias correction (Begg–Greenes method) for EU-TIRADS
- Supplementary Table S12. Algorithmic fairness audit — disparate impact ratio at the Youden threshold (sealed test set)
- Supplementary Table S13. Spectrum sensitivity analysis — reinstating the excluded indeterminate nodules
- Section F. Supplementary figures
- Supplementary Figure S1. EU-TIRADS categories in the analyzed cohort
- Supplementary Figure S2. Malignancy rate by the number of concurrent high-suspicion sonographic features
- Supplementary Figure S3. Distribution of predicted malignancy probabilities by reference-standard status

#### Section A. Reporting checklists

##### Supplementary Table S1. STARD 2015 checklist

| # | Item | Where reported |
|---|---|---|
| 1 | Identification as a study of diagnostic accuracy | Title; Abstract |
| 2 | Structured summary | Abstract |
| 3 | Scientific and clinical background | Introduction, paragraphs 1–3 |
| 4 | Study objectives and hypotheses | Introduction, final paragraph; Materials and Methods, “Hypotheses” |
| 5 | Prospective or retrospective collection | Materials and Methods, “Design, setting and reporting” (retrospective) |
| 6 | Eligibility criteria | Materials and Methods, “Participants and sources” |
| 7–8 | On what basis, where and when participants were identified | Materials and Methods, “Participants and sources” (consecutive recruitment in the two Tunisian centers; selection from a public dataset) |
| 9 | Whether participants formed a consecutive, random or convenience series | Materials and Methods, “Design” (consecutive in the two Tunisian centers; the 103 public-dataset nodules are a selection) |
| 10a | Index test, in sufficient detail | Materials and Methods; Table 1; Section C |
| 10b | Reference standard, in sufficient detail | Materials and Methods, “Reference standard” |
| 11 | Rationale for choosing the reference standard | Materials and Methods, “Reference standard” (two-tier standard; verification-bias rationale) |
| 12a | Definition of and rationale for index test positivity cut-offs | Materials and Methods, “Statistical analysis”; Table 3 (three thresholds); Supplementary Table S8 (threshold grid) |
| 12b | Definition of and rationale for reference standard positivity cut-offs | Materials and Methods, “Reference standard” (Bethesda II vs V/VI; histology when available) |
| 13a | Whether clinical information was available to the index test performers | Materials and Methods, “Model, fine-tuning and configurations” (inputs enumerated; EU-TIRADS withheld) |
| 13b | Whether clinical information was available to the reference standard assessors | Materials and Methods, “Participants and sources” (reference standard established in routine care, before and independently of the model) |
| 14 | Methods for estimating or comparing measures of diagnostic accuracy | Materials and Methods, “Statistical analysis” |
| 15 | How indeterminate index test or reference standard results were handled | Materials and Methods, “Reference standard”; Figure 1; Supplementary Table S13 |
| 16 | How missing data on the index test and reference standard were handled | Materials and Methods, “Data partition and sealing”; Results, “Cohort”; Supplementary Table S11 |
| 17 | Any analyses of variability in diagnostic accuracy | Materials and Methods, “Statistical analysis”; Figure 6; Supplementary Tables S10 and S12 |
| 18 | Intended sample size and how it was determined | Materials and Methods, “Sample size” |
| 19 | Flow of participants | Figure 1 |
| 20 | Baseline demographic and clinical characteristics | Table 2 |
| 21a | Distribution of severity of disease in those with the target condition | Table 2 (EU-TIRADS, Bethesda, size); Supplementary Figure S1 |
| 21b | Distribution of alternative diagnoses in those without the target condition | Table 2 |
| 22 | Time interval and any clinical interventions between index test and reference standard | Materials and Methods, “Data partition and sealing”: interval not recorded (year of examination only) |
| 23 | Cross-tabulation of index test results by the reference standard | Table 3; Figure 4A–B |
| 24 | Estimates of diagnostic accuracy and their precision | Table 3; Figure 3 |
| 25 | Any adverse events | Not applicable (retrospective analysis of existing images and reports) |
| 26 | Study limitations | Discussion, “Limitations” |
| 27 | Implications for practice | Discussion, “Clinical implications” |
| 28 | Registration number and name of registry | Not registered; ethics approval CEM-2025-10-04 |
| 29 | Where the full study protocol can be accessed | No protocol published (Declarations); split definition available from the corresponding author |
| 30 | Sources of funding and other support; role of funders | Declarations, “Funding” (none) |

##### Supplementary Table S2. TRIPOD+AI checklist (key items)

| Domain | Item | Where reported |
|---|---|---|
| Title and abstract | Model type, target population, outcome, intended use | Title; Abstract |
| Introduction | Rationale, existing models, objectives | Introduction |
| Methods — data | Data sources, eligibility, dates, setting | Materials and Methods, “Participants and sources” |

| Domain | Item | Where reported |
|---|---|---|
| Methods — data | Handling of the train/validation/test split, and safeguards against leakage | Materials and Methods, “Data partition and sealing”; Figure 1 |
| Methods — outcome | Definition, blinding of outcome assessment | Materials and Methods, “Reference standard” |
| Methods — predictors | Definition, timing, blinding; predictors deliberately withheld | Materials and Methods, “Model, fine-tuning and configurations”; Section B |
| Methods — sample size | Justification | Materials and Methods, “Sample size” |
| Methods — missing data | Approach | Materials and Methods; Supplementary Table S11 |
| Methods — model | Architecture, fine-tuning, hyperparameters, seeds, hardware | Materials and Methods; Section C |
| Methods — model | Class imbalance handling | Materials and Methods (class-weighted cross-entropy) |
| Methods — evaluation | Discrimination, calibration, clinical utility; comparator | Materials and Methods, “Statistical analysis” |
| Methods — fairness | Subgroup and equity analyses planned | Materials and Methods; Supplementary Tables S10 and S12 |
| Results — participants | Flow, characteristics, outcome frequency | Figure 1; Table 2 |
| Results — model performance | Point estimates with uncertainty at all evaluated thresholds | Tables 3–7; Figures 3–5; Supplementary Table S8 |
| Results — model updating | Recalibration | Table 7; Figure 5B; Supplementary Table S9 |
| Discussion | Interpretation, limitations, usability, generalisability | Discussion |
| Other | Funding, conflicts, data and code availability, AI-tool use | Declarations |

#### Supplementary Table S3. CLAIM 2024 checklist (key items)

| # | Item | Where reported |
|---|---|---|
| 1–2 | Identification as an AI study; structured summary | Title; Abstract |
| 3–5 | Background, objectives, study design | Introduction; Materials and Methods |
| 6–9 | Data sources, eligibility, data pre-processing, de-identification | Materials and Methods, “Participants and sources”; “Image acquisition and pre-processing” |
| 10–12 | Ground truth definition, rater qualifications, measurement of agreement | Materials and Methods, “Reference standard”; “Participants and sources” (30, at least 20 and 5 years’ experience, by source) |
| 13–15 | Partitions: how and when defined, level of partitioning, leakage prevention | Materials and Methods, “Data partition and sealing” (patient level, sealed, SHA-256) |
| 16–19 | Model description, initialization, training approach, hyperparameters | Materials and Methods; Table 1; Section C |
| 20–22 | Software, hardware, randomization control | Materials and Methods; Section C (seed 42, bfloat16, RTX PRO 6000 and A100) |
| 23–26 | Evaluation metrics, statistical measures of significance and uncertainty, robustness, failure analysis | Materials and Methods, “Statistical analysis”; Tables 3–7; Supplementary Tables S10–S13 |
| 27–30 | Results: flow, demographics, performance, comparison with a reference | Figure 1; Tables 2–7 |
| 31–33 | Discussion: limitations, implications, model and data availability | Discussion; Declarations |

### Section B. Variables and prompt construction

#### Supplementary Table S4. Variables collected and their use as model inputs

| Variable | Label | Type | Levels / unit | Supplied to the model? |
|---|---|---|---|---|
| ID_NOD | Nodule identifier | ID | pseudonymized code | No |
| CTR_ID | Recruitment source | categorical | 1 = Mahdia; 2 = international public dataset; 3 = Tunis | No |
| AGE | Age at FNA | numeric | years | Configs A, C, D |
| SEX | Sex | binary | 1 = male; 2 = female | Configs A, C, D |
| EU_TIRADS | EU-TIRADS category | ordinal | 2, 3, 4, 5 | **Never** (comparator) |
| COMP | Composition | categorical | solid / mixed / cystic | Config D |
| Spongiforme | Spongiform appearance | binary | 0 = no; 1 = yes | Config D |
| ECHOG | Echogenicity | categorical | 1 = hyperechoic; 2 = isoechoic; 3 = hypoechoic; 4 = | Config D |

| Variable | Label | Type | Levels / unit | Supplied to the model? |
|---|---|---|---|---|
| | | | markedly hypoechoic | |
| SHAPE | Shape | binary | 0 = oval/round; 1 = taller than wide | Config D |
| MARG | Margins | categorical | 1 = smooth; 2 = angular (irregular in EU-TIRADS terms); 3 = ill-defined; 4 = lobulated | Config D |
| MICROCALC | Microcalcifications | binary | 0 = absent; 1 = present | Config D |
| MACROCAL_C | Central macrocalcifications | binary | 0 = absent; 1 = present | Config D |
| MACROCAL_PER | Peripheral macrocalcifications | binary | 0 = absent; 1 = present | Config D |
| Halo | Perinodular halo | categorical | 0 = absent; 1 = thin/complete; 3 = interrupted | Config D |
| VASC_CENT | Central vascularity (color Doppler, radiologist's report) | binary | 0 = absent; 1 = present | Config D |
| CONTACT_CAPS | Capsular contact | binary | 0 = absent; 1 = present | Config D |
| PUNCT_ECHOG | Echogenic foci | categorical | 0 = absent; 1 = colloid (comet-tail); 2 = sediment-like; 3 = microcystic | Config D |
| D1_mm, D2_mm, D3_mm | Orthogonal diameters | numeric | mm | Configs A, B, C, D |
| MAXD_mm | Maximum diameter | numeric | mm | Configs A, B, C, D |
| VOL_cc | Nodule volume (ellipsoid formula) | numeric | $cm^3$ | Configs A, B, C, D |
| Volume_T | Thyroid volume | numeric | $cm^3$ | No |
| EMPL | Vertical location (not supplied to the model) | categorical | 1 = upper pole; 2 = mid-gland; 3 = lower pole | No |
| RAD_EXP | Experience of the rating radiologist | numeric | years | No |
| US_MFR, US_MODEL | Scanner manufacturer and model | categorical | — | **No** (acquisition metadata without semiological content) |
| BETHESDA | Bethesda cytology category (Tunisian centers; not applicable to the public dataset, whose reference is histological) | ordinal | II–VI | **No** (label leakage) |
| HISTO_DX | Surgical histological diagnosis | text | — | **No** (label leakage) |
| OUTCOME_SRC | Source of the outcome | categorical | 1 = cytology; 2 = histology | No |
| OUTCOME_BIN | Reference standard | binary | 0 = benign; 1 = malignant | Target |
| SPLIT | Canonical partition | categorical | train / validation / test / excluded | — |

Three families of variables were deliberately withheld from every configuration: the scanner model (acquisition metadata carrying no semiological information), free-text cytological description (unnormalized string), and the Bethesda category (the label itself). The overall EU-TIRADS category was likewise withheld, in order to preserve the independence of the human comparator. For the public dataset, the EU-TIRADS category, the nodule measurements and the eleven qualitative descriptors were coded by one of two radiologists with 5 years' experience, blinded to histology.

#### B.1 Prompt construction

Tabular variables were serialized as structured natural-language text inside the user turn, immediately after the image token, in a fixed field order. Fields were rendered as name: value pairs with units, using the human-readable level names of the table above rather than numeric codes. No free text, no derived score and no textual hint about malignancy was inserted. The five biometric–demographic prompt fields are age, sex, nodule volume, dimensions (three orthogonal and maximum diameters) and size-derived ratios; among the eleven descriptor fields, central and peripheral macrocalcifications form a single field. No descriptor was missing: the 16 input fields were complete for all 1,456 included nodules, in every partition, so no imputation was required. The system turn framed the task as binary risk estimation and requested a probability; the assistant turn during fine-tuning contained the target label together with the auxiliary sonographic targets.

#### Section C. Training protocol

##### Supplementary Table S5. Shared training configuration

| Element | Value |
|---|---|
| Base model | MedGemma 1.5 4B (google/medgemma-1.5-4b-it) |
| Vision encoder | MedSigLIP (SigLIP ViT-So400m), input 896 × 896, patch 14 × 14 |
| Adaptation | LoRA, rank r = 64, α = 64 (α/r = 1.0), dropout 0.05 |
| LoRA targets | all linear layers (all-linear), multimodal projector included |
| Trainable parameters | ≈157 million (≈3.5% of the total parameters of the adapted model); base weights frozen |
| Optimiser | AdamW, weight decay 0.01 |
| Learning rate | $2 \times 10^{-4}$, cosine schedule, 10% warm-up (5% for Config C) |
| Gradient clipping | $\|\nabla\| \leq 1.0$ |
| Effective batch size | 16 (micro-batch 4 × gradient accumulation 4) |
| Sequence length | 768 tokens (Configs A, B); 1,024 tokens (Configs C, D) |
| Epochs | ≤ 10; validation every 50 optimization steps, early stopping with patience 5 evaluations |
| Loss | class-weighted cross-entropy (benign 0.6149, malignant 2.6765, i.e. n/(2·n_class) on the 1,456 analyzed nodules; training partition alone: 0.609 and 2.787); class-weighted sampling of training examples (malignant weight 5.0) |
| Precision | bfloat16; PyTorch 2.8.0 (Configs A, B) and 2.10.0 (Configs C, D), CUDA 12.8; SDPA attention with FlashAttention-2 |
| Hardware | local servers: NVIDIA RTX PRO 6000 (Configs A, B) and NVIDIA A100-SXM4 80 GB (Configs C, D) |
| Random seed | 42 (initialization, shuffling, augmentation, all analyses) |

Class weighting (of the loss and of the sampling of training examples) was preferred to synthetic oversampling (SMOTE-type), which creates artificial examples and can bias latent representations, and to focal loss, because it preserves the original data distribution while remaining simple and transparent.

##### C.1 Data augmentation

Applied on the fly, each with probability 0.5: horizontal flip; photometric adjustment (± 15%); Gaussian blur ($\sigma \in [0.1, 1.0]$); restricted affine rotation (± 10°). Elastic deformation and vertical flip were **not** permitted, as either would alter the semiology of shape and margins on which the diagnosis rests. No destructive smoothing filter was applied at pre-processing, in order to preserve native speckle texture.

##### Supplementary Table S6. Differences between the four configurations

| Configuration | Inputs beyond the image | Auxiliary EU-TIRADS target | Other |
|---|---|---|---|
| Config A | nodule biometry (D1, D2, D3, Dmax, volume) + age and sex | yes | reference multimodal configuration |
| Config B | nodule biometry alone | yes | ablation isolating nodule biometry |
| Config C | as Config A | **no** | corrected logit extraction; chain-of-thought prompting with five ground-truth-supervised semiological steps; five auxiliary sonographic targets in place of the EU-TIRADS target |
| Config D | the inputs of Config A plus the 11 qualitative descriptors of the structured report (16 prompt fields in all) | yes | full qualitative sonographic description; random masking of report fields during training (0.10–0.15 per field; all fields in 10% of examples) |

Configurations A, B and C require no qualitative interpretation from the radiologist: they consume the image and the nodule dimensions measured during any routine examination, with or without the basic patient characteristics available on a request form. Config D is the only configuration that consumes the radiologist's structured interpretation. The exact variable-by-variable mapping is given in Supplementary Table S4, which is the formal predictor specification required by TRIPOD+AI.

#### C.2 Development of the configurations

The four configurations were defined to isolate the contribution of each family of inputs: nodule biometry with or without age and sex (Configs A and B), supervision of intermediate sonographic reasoning (Config C) and the radiologist's qualitative descriptors (Config D). Preliminary runs, used to stabilize logit extraction and the prompt format, were not evaluated on the test set. The four configurations were evaluated once, on the same locked test split.

#### Supplementary Table S7. Checkpoint selection and validation performance

| Configuration | Epoch at early stopping | Best checkpoint (epoch) | Validation AUROC | Validation AUPRC | Validation sensitivity |
|---|---|---|---|---|---|
| Config A | 5.89 | 3.43 | 0.817 | 0.553 | 63.5% |
| Config B | 5.40 | 2.95 | 0.803 | 0.595 | 51.9% |
| Config C | 4.41 | 1.97 | 0.779 | 0.492 | 51.9% |
| Config D | 6.38 | 3.93 | **0.908** | **0.694** | **78.8%** |

Validation set n = 258 (52 malignant, 20.2%). The checkpoint retained for each configuration is the one with the best validation performance under early stopping (patience 5 evaluations). Validation loss plateaued and validation AUROC saturated well before the tenth planned epoch in every configuration, so the effective training length was 2–4 epochs. These quantities come from the training logs; unlike every other value in this article they could not be recomputed from a persisted per-sample prediction file, because validation probabilities were not retained. The ordering of configurations on the validation set is the same as on the sealed test set; this concordance is not independent evidence, since the validation set governed checkpoint selection.

### Section D. Methodological definitions

**AUROC.** Area under the receiver operating characteristic curve; the probability that a randomly chosen malignant nodule receives a higher predicted score than a randomly chosen benign one. Invariant to any monotone transformation of the score, and therefore unaffected by recalibration or by the particular category-to-probability mapping used for EU-TIRADS.

**EU-TIRADS probability mapping.** To compute threshold-free metrics, the EU-TIRADS categories were mapped to working probabilities fixed by the authors before the analysis: T2 → 0.02, T3 → 0.07, T4 → 0.20, T5 → 0.55. The value for category 5 follows the mean risk reported by the European consensus for nodules with high-risk features (Russ et al., Eur Thyroid J 2017; doi:10.1159/000478927); the values for categories 3 and 4 lie within the 95% confidence intervals of the pooled malignancy prevalences of a meta-analysis of EU-TIRADS (Castellana et al., Eur J Endocrinol 2020; doi:10.1530/EJE-20-0204); category 2 corresponds to a risk close to zero. These are not published point estimates. The mapping leaves AUROC and the DeLong comparisons unchanged; it determines only the Brier score, the calibration metrics and the net benefit of EU-TIRADS, and the coincidence of the 20% decision-curve threshold with the probability of category 4 follows from it.

**AUPRC.** Area under the precision–recall curve. Preferable to AUROC under class imbalance because it is unaffected by the large number of true negatives; the no-skill reference is the prevalence (0.199 here), not 0.5.

**BCa bootstrap.** Bias-corrected and accelerated bootstrap confidence interval; corrects both for median bias and for skewness of the bootstrap distribution, which matters for statistics bounded at 1 such as AUROC.

**DeLong test.** Non-parametric comparison of two AUROCs estimated on the *same* subjects, accounting for their correlation through the structural components of the Mann–Whitney statistic. Appropriate here because model and comparator are evaluated on identical nodules.

**Youden index.** J = sensitivity + specificity − 1, maximized over thresholds. Derived on the evaluation set itself, the corresponding threshold is an optimistic upper bound of the achievable trade-off; derived on the validation partition, it can be carried unchanged to new data.

**Brier score.** Mean squared difference between predicted probability and observed outcome; 0 is perfect. **Brier skill score** = 1 − (Brier / Brier of a prevalence-only predictor); values below 0 indicate probabilities less informative than simply quoting the prevalence.

**Expected calibration error (ECE).** Weighted mean absolute difference between predicted probability and observed frequency across bins of predicted probability (10 equal-width bins here). Sensitive to the binning scheme, which is why it is reported alongside calibration-in-the-large and slope.

**Calibration-in-the-large and calibration slope.** Intercept of a logistic recalibration of the outcome on the logit of the predicted probability with the slope fixed at 1 (ideal 0), and the free slope of the same recalibration (ideal 1; > 1 indicates under-confidence, < 1 over-confidence).

**Spiegelhalter's z test.** Global test of calibration comparing the observed and expected mean squared error under the null hypothesis of perfect calibration.

**Platt scaling.** Recalibration fitting a one-parameter logistic map on the logit of the predicted probability. Monotone, therefore leaves the ranking — and hence AUROC — unchanged.

**Net reclassification improvement (binary).** (Correctly reclassified events − incorrectly reclassified events) / number of events, plus the same quantity among non-events. Reported here against EU-TIRADS ≥ 4, so it measures how the model redistributes nodules relative to the clinically used comparator rather than relative to chance.

**Decision curve analysis / net benefit.** NB = (true positives / n) − (false positives / n) × [t / (1 − t)], where t is the threshold probability at which a clinician would choose to biopsy. It makes explicit the exchange rate between a missed cancer and an unnecessary aspiration, and allows comparison with the two reference strategies "biopsy all" and "biopsy none".

**Disparate impact ratio.** Ratio of the positive prediction rate within a stratum to the positive prediction rate in the whole evaluation set. The conventional acceptable band, 0.80–1.25, is the bilateral extension of the four-fifths rule. It measures differential *rates of positive calls*, not differential *error rates*.

**Verification (work-up) bias and the Begg–Greenes correction.** Bias arising when the reference standard is obtained preferentially in test-positive subjects. The correction reweights verified counts by the conditional probability of verification within each level of the test, under a conditional missing-at-random assumption.

**Spectrum effect.** Variation of sensitivity, specificity and predictive values with the case-mix of the population tested. Relevant here because the cohort consists exclusively of aspirated nodules and excludes cytologically indeterminate ones.

---

#### Section E. Supplementary results

##### Supplementary Table S8. Config D across decision thresholds: cancers detected and aspirations indicated (sealed test set)

| Threshold | Called positive | Cancers detected (of 51) | Cancers missed | Benign called positive (of 205) | Se, % | Sp, % | Δ aspirations vs EU-TIRADS ≥ 4 | Δ cancers missed vs EU-TIRADS ≥ 4 |
|---|---|---|---|---|---|---|---|---|

| Threshold | Called positive | Cancers detected (of 51) | Cancers missed | Benign called positive (of 205) | Se, % | Sp, % | Δ aspirations vs EU-TIRADS ≥ 4 | Δ cancers missed vs EU-TIRADS ≥ 4 |
|---|---|---|---|---|---|---|---|---|
| EU-TIRADS ≥ 4 (comparator) | 144 | 50 | 1 | 94 | 98.0 | 54.1 | reference | reference |
| 0.05 | 231 | 51 | 0 | 180 | 100.0 | 12.2 | +87 | −1 |
| 0.10 | 150 | 50 | 1 | 100 | 98.0 | 51.2 | +6 | 0 |
| 0.15 | 121 | 49 | 2 | 72 | 96.1 | 64.9 | −23 | +1 |
| 0.20 | 108 | 49 | 2 | 59 | 96.1 | 71.2 | −36 | +1 |
| 0.25 | 96 | 49 | 2 | 47 | 96.1 | 77.1 | −48 | +1 |
| 0.30 | 77 | 49 | 2 | 28 | 96.1 | 86.3 | −67 | +1 |
| 0.3208 (Youden, test set) | 74 | 49 | 2 | 25 | 96.1 | 87.8 | −70 | +1 |
| 0.3346 (Youden, validation partition) | 70 | 48 | 3 | 22 | 94.1 | 89.3 | −74 | +2 |
| 0.40 | 57 | 43 | 8 | 14 | 84.3 | 93.2 | −87 | +7 |
| 0.50 | 41 | 34 | 17 | 7 | 66.7 | 96.6 | −103 | +16 |
| 0.60 | 22 | 20 | 31 | 2 | 39.2 | 99.0 | −122 | +30 |

Config D on the sealed test set (n = 256; 51 malignant, 205 benign). A nodule is called positive, that is, referred for aspiration, when its predicted probability is greater than or equal to the threshold. The last two columns compare each threshold with EU-TIRADS ≥ 4 (144 aspirations, 1 cancer missed): negative values are aspirations spared or fewer cancers missed, positive values additional aspirations or additional cancers missed. Because the predicted probabilities take discrete values, neighboring thresholds can give identical counts. Se, sensitivity; Sp, specificity. The thresholds of 0.3208 and 0.3346 are the Youden thresholds of the test set and of the validation partition (Table 3). The native output of the model is a generated label rather than a probability threshold; it is reported in Table 3.

---

#### Supplementary Table S9. Recalibration by Platt scaling (internal cross-validation on the sealed test set)

| Configuration | ECE raw → Platt | Brier raw → Platt | Observed/expected raw → Platt | AUROC raw → Platt |
|---|---|---|---|---|
| Config A | 0.192 → **0.032** | 0.191 → 0.101 | 29.37 → 1.00 | 0.892 → 0.888 |
| Config B | 0.195 → **0.026** | 0.194 → 0.098 | 43.17 → 0.99 | 0.886 → 0.881 |
| Config C | 0.114 → **0.054** | 0.113 → 0.098 | 1.59 → 1.00 | 0.883 → 0.877 |
| Config D | 0.117 → **0.019** | 0.080 → 0.063 | 0.84 → 1.00 | 0.953 → 0.948 |

Because per-sample predicted probabilities were persisted for the sealed test set only, recalibration could not be fitted on the independent validation set. It was therefore estimated by 5-fold stratified cross-validation within the test set (seed 42, out-of-fold predictions), which quantifies the correctability of the miscalibration without providing a deployable recalibration map. Platt (sigmoid) scaling is reported; isotonic regression achieved comparable expected calibration error but overfitted at this sample size (slope ≈ 0.43 with slight AUROC erosion). Because recalibration is a monotone transformation, AUROC is preserved up to cross-validation variance (≤ 0.006). Caveat: the post-recalibration calibration error depends on the fold draw. Across ten seeds, Config D ranges from 0.019 to 0.040 (median 0.031); the value reported here corresponds to seed 42, declared a priori in the Materials and Methods, and lies at the low end of that range. The conclusion — a calibration deficit correctable without loss of discrimination — holds for every seed, the raw ECE being 0.117; the figure of 0.019 should nonetheless not be read as a stable point estimate.

---

#### Supplementary Table S10. AUROC by prespecified subgroup, all four configurations (sealed test set)

| Stratum | n | Malignant | Config A | Config B | Config C | Config D |
|---|---|---|---|---|---|---|
| **Recruitment source** | | | | | | |

| Stratum | n | Malignant | Config A | Config B | Config C | Config D |
|---|---|---|---|---|---|---|
| Mahdia † | 42 | 6 | 0.769 (0.562–0.922) | 0.745 (0.433–0.986) | 0.778 (0.523–0.944) | 0.963 (0.857–1.000) |
| Public dataset † | 46 | 4 | 0.863 (0.679–0.964) | 0.878 (0.726–0.964) | 0.893 (0.611–1.000) | 0.994 (0.913–1.000) |
| Tunis | 168 | 41 | 0.916 (0.865–0.952) | 0.905 (0.832–0.946) | 0.904 (0.847–0.942) | 0.949 (0.900–0.975) |
| **Maximum diameter** | | | | | | |
| <10 mm † | 24 | 19 | 0.632 (0.200–0.916) | 0.858 (0.657–0.963) | 0.611 (0.242–0.895) | 0.647 (0.347–0.905) |
| 10–20 mm | 79 | 27 | 0.709 (0.581–0.816) | 0.693 (0.570–0.802) | 0.667 (0.533–0.779) | 0.892 (0.792–0.950) |
| >20 mm † | 153 | 5 | 0.803 (0.614–0.938) | 0.749 (0.463–0.981) | 0.682 (0.380–0.906) | 0.992 (0.958–1.000) |
| **Age** | | | | | | |
| <55 years | 145 | 32 | 0.848 (0.770–0.902) | 0.873 (0.789–0.926) | 0.839 (0.740–0.901) | 0.948 (0.886–0.977) |
| ≥55 years | 111 | 19 | 0.939 (0.862–0.978) | 0.905 (0.765–0.965) | 0.946 (0.877–0.982) | 0.959 (0.875–0.989) |
| **Sex** | | | | | | |
| Men † | 29 | 6 | 0.899 (0.710–0.978) | 0.906 (0.707–1.000) | 0.873 (0.642–1.000) | 0.935 (0.768–0.993) |
| Women | 227 | 45 | 0.893 (0.839–0.932) | 0.883 (0.813–0.929) | 0.883 (0.807–0.924) | 0.953 (0.906–0.977) |
| **EU-TIRADS category** | | | | | | |
| EU-TIRADS 3 † | 111 | 1 | 0.673 (0.577–0.750) | 0.286 (0.205–0.364) | 0.855 (0.773–0.909) | 0.768 (0.682–0.836) |
| EU-TIRADS 4 † | 87 | 3 | 0.925 (0.774–0.988) | 0.851 (0.589–0.982) | 0.849 (0.670–0.956) | 0.631 (0.196–0.869) |
| EU-TIRADS 5 | 57 | 47 | 0.528 (0.330–0.740) | 0.635 (0.478–0.792) | 0.510 (0.306–0.736) | 0.630 (0.417–0.800) |

95% bias-corrected and accelerated bootstrap intervals stratified on outcome (B = 2,000, seed 42). † fewer than 10 events or fewer than 10 non-events in the stratum; the estimate is not interpretable. Strata with fewer than 10 nodules, or with a single outcome class, were not computed.

#### Supplementary Table S11. Verification-bias correction (Begg–Greenes method) for EU-TIRADS

##### (a) Verification rate by EU-TIRADS category (verification population, n = 1,560: the 20 public-dataset images excluded for image reasons are not unverified nodules)

| EU-TIRADS category | Eligible nodules | Verified nodules | P(verified \| category) | P(malignant \| category, verified) |
|---|---|---|---|---|
| 2 | 8 | 8 | 1.000 | 0.000 |
| 3 | 609 | 597 | 0.980 | 0.012 |
| 4 | 631 | 562 | 0.891 | 0.068 |
| 5 | 312 | 289 | 0.926 | 0.785 |
| **Total** | 1,560 | 1,456 | 0.933 | — |

##### (b) Observed versus verification-bias–corrected accuracy

| Threshold | Sensitivity observed | Sensitivity corrected | Specificity observed | Specificity corrected |
|---|---|---|---|---|
| EU-TIRADS ≥ 4 | 97.4% | 97.6% | 50.5% | 48.2% |
| EU-TIRADS ≥ 5 | 83.5% | 83.1% | 94.8% | 94.7% |

Reweighting of the verified counts by the conditional probability of verification within each EU-TIRADS category. Unverified nodules concentrate in category 4 (69 of 631, 10.9%), which is precisely where the ≥ 4 threshold operates; correction therefore moves specificity more than sensitivity. The conditional missing-at-random assumption — that the decision to obtain a definitive reference standard does not depend on true malignancy given the EU-TIRADS category — is debatable here, because histological verification follows the surgical decision.

#### Supplementary Table S12. Algorithmic fairness audit — disparate impact ratio at the Youden threshold (sealed test set)

| Configuration | Variable | Stratum | n | Positive rate | Disparate impact ratio | Outside 0.80–1.25 |
|---|---|---|---|---|---|---|
| Config A | Sex | Men | 29 | 0.517 | 1.39 | **yes** |
| | Sex | Women | 227 | 0.352 | 0.95 | no |
| | Recruitment source | Mahdia | 42 | 0.167 | 0.45 | **yes** |
| | Recruitment source | Public dataset | 46 | 0.370 | 1.00 | no |
| | Recruitment source | Tunis | 168 | 0.423 | 1.14 | no |
| | Age band | <30 y | 13 | 0.385 | 1.04 | no |
| | Age band | 30-39 y | 45 | 0.422 | 1.14 | no |
| | Age band | 40-49 y | 65 | 0.462 | 1.24 | no |
| | Age band | 50-59 y | 47 | 0.383 | 1.03 | no |
| | Age band | 60-69 y | 54 | 0.278 | 0.75 | **yes** |
| | Age band | ≥70 y | 32 | 0.250 | 0.67 | **yes** |
| Config B | Sex | Men | 29 | 0.379 | 1.18 | no |
| | Sex | Women | 227 | 0.313 | 0.98 | no |
| | Recruitment source | Mahdia | 42 | 0.190 | 0.59 | **yes** |
| | Recruitment source | Public dataset | 46 | 0.348 | 1.09 | no |
| | Recruitment source | Tunis | 168 | 0.345 | 1.08 | no |
| | Age band | <30 y | 13 | 0.385 | 1.20 | no |
| | Age band | 30-39 y | 45 | 0.378 | 1.18 | no |
| | Age band | 40-49 y | 65 | 0.308 | 0.96 | no |
| | Age band | 50-59 y | 47 | 0.362 | 1.13 | no |
| | Age band | 60-69 y | 54 | 0.222 | 0.69 | **yes** |
| | Age band | ≥70 y | 32 | 0.344 | 1.07 | no |
| Config C | Sex | Men | 29 | 0.448 | 1.33 | **yes** |
| | Sex | Women | 227 | 0.322 | 0.96 | no |
| | Recruitment source | Mahdia | 42 | 0.190 | 0.57 | **yes** |
| | Recruitment source | Public dataset | 46 | 0.348 | 1.04 | no |
| | Recruitment source | Tunis | 168 | 0.369 | 1.10 | no |
| | Age band | <30 y | 13 | 0.231 | 0.69 | **yes** |
| | Age band | 30-39 y | 45 | 0.422 | 1.26 | **yes** |
| | Age band | 40-49 y | 65 | 0.446 | 1.33 | **yes** |
| | Age band | 50-59 y | 47 | 0.277 | 0.82 | no |
| | Age band | 60-69 y | 54 | 0.259 | 0.77 | **yes** |
| | Age band | ≥70 y | 32 | 0.250 | 0.74 | **yes** |
| Config D | Sex | Men | 29 | 0.414 | 1.43 | **yes** |
| | Sex | Women | 227 | 0.273 | 0.94 | no |
| | Recruitment source | Mahdia | 42 | 0.214 | 0.74 | **yes** |
| | Recruitment source | Public dataset | 46 | 0.196 | 0.68 | **yes** |
| | Recruitment source | Tunis | 168 | 0.333 | 1.15 | no |
| | Age band | <30 y | 13 | 0.231 | 0.80 | **yes** |
| | Age band | 30-39 y | 45 | 0.378 | 1.31 | **yes** |
| | Age band | 40-49 y | 65 | 0.415 | 1.44 | **yes** |
| | Age band | 50-59 y | 47 | 0.298 | 1.03 | no |
| | Age band | 60-69 y | 54 | 0.148 | 0.51 | **yes** |
| | Age band | ≥70 y | 32 | 0.156 | 0.54 | **yes** |

Disparate impact ratio = (positive prediction rate within the stratum) / (positive prediction rate in the whole test set), evaluated at each configuration's Youden threshold. The acceptable range is the bilateral extension of the Equal Employment Opportunity Commission four-fifths rule (0.80–1.25). Strata with fewer than 10 nodules are omitted. A ratio outside the range signals a differential rate of positive calls, not necessarily a differential error rate; subgroup AUROCs are given in Supplementary Table S10.

#### Supplementary Table S13. Spectrum sensitivity analysis — reinstating the excluded indeterminate nodules

| Population | n | Ordinal AUROC of EU-TIRADS |
|---|---|---|
| Analyzed cohort (indeterminate nodules excluded) | 1,456 | 0.921 |
| Plus 104 Tunisian Bethesda III–IV nodules, all assumed benign | 1,560 | 0.912 |
| Plus 104 Tunisian Bethesda III–IV nodules, all assumed malignant | 1,560 | 0.867 |

Worst-case reclassification analysis. Excluding the cytologically indeterminate nodules raises the apparent accuracy of EU-TIRADS in the analyzed cohort; the malignant-assumption scenario is the pessimistic bound and brings the estimate closer to what is reported in less selected series of aspirated nodules. The same spectrum effect applies to the model, which was trained and evaluated on the same distribution.

### Section F. Supplementary figures

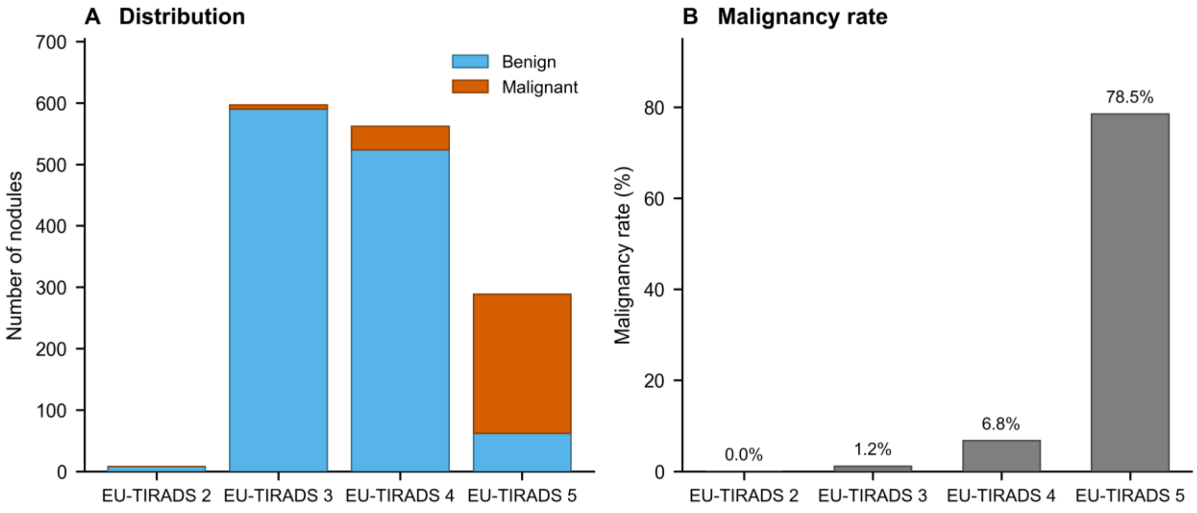


**Supplementary Figure S1. EU-TIRADS categories in the analyzed cohort (n = 1,456).** (A) Absolute numbers of benign and malignant nodules per category. (B) Malignancy rate per category, rising monotonically from 0% (EU-TIRADS 2) to 78.5% (EU-TIRADS 5) (Cochran–Armitage trend $p < 0.001$). The cohort is dominated by intermediate categories 3 and 4, which together account for 79.6% of nodules but only 16.5% of cancers.

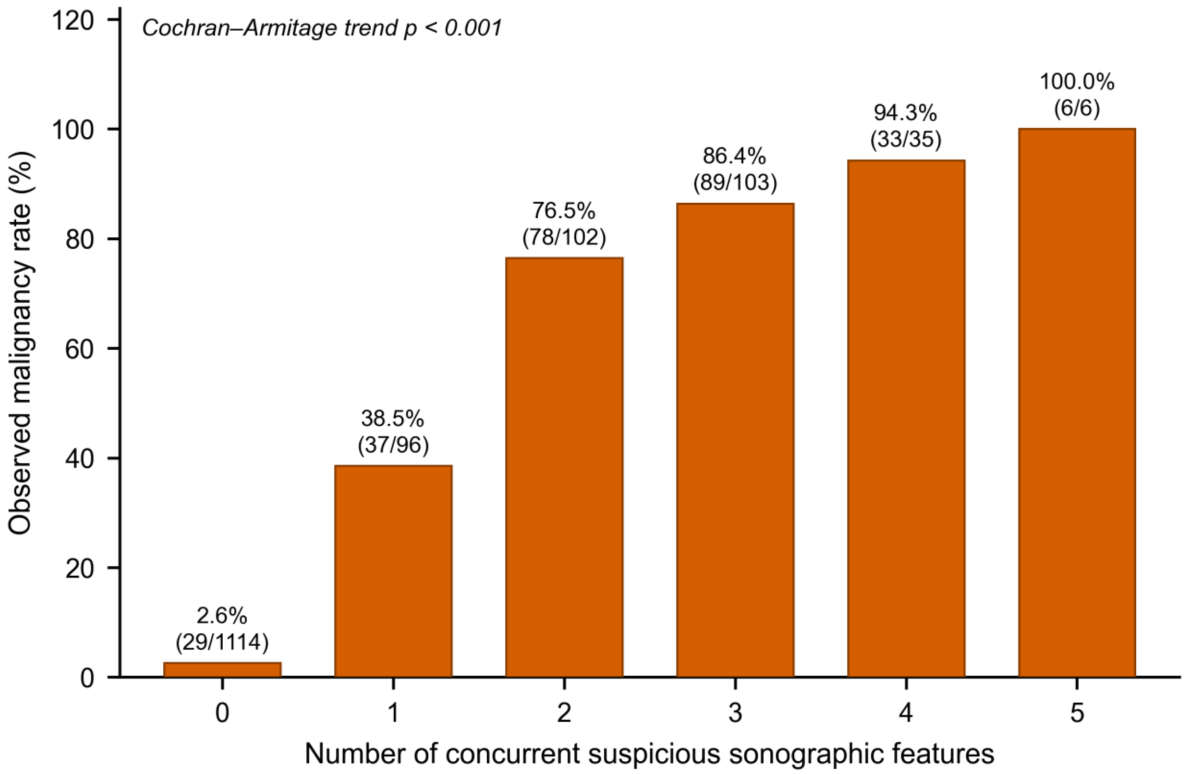


**Supplementary Figure S2. Malignancy rate by the number of concurrent high-suspicion sonographic features (analyzed cohort, n = 1,456).** Features counted are the four EU-TIRADS high-suspicion signs — markedly hypoechoic parenchyma, irregular (angular, ill-defined or lobulated) margins, microcalcifications and taller-than-wide shape — plus capsular contact as an additional descriptor. The malignancy rate rises monotonically from 2.6% with no feature to 100% with all five (Cochran–Armitage trend $p < 0.001$). Note that only marked hypoechogenicity qualifies as a high-suspicion sign under EU-TIRADS; counting simple hypoechogenicity as well would misclassify a large fraction of benign nodules as carrying a suspicious feature.

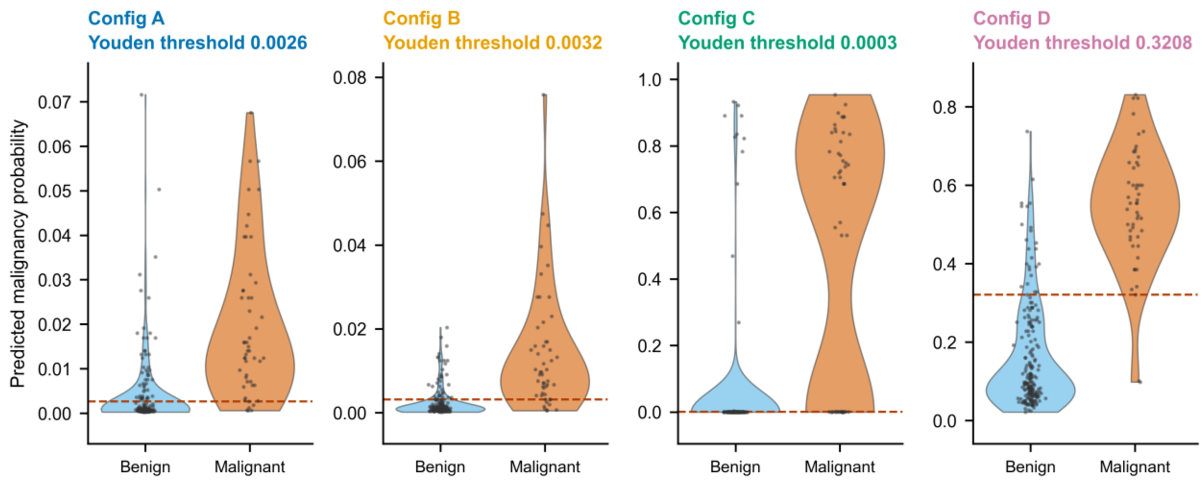


**Supplementary Figure S3. Distribution of predicted malignancy probabilities by reference-standard status (sealed test set, n = 256).** Violin plots with individual nodules overlaid; benign in blue, malignant in orange. The dashed line marks each configuration's Youden threshold. Configs A, B and C compress virtually all probability mass below 0.08 — hence Youden thresholds of the order of $10^{-3}$ — whereas Config D spans a usable dynamic range with a threshold of 0.321. Mann–Whitney U test $p < 0.001$ for all four configurations. Ranking (AUROC) and the probability scale (calibration, net benefit) are therefore distinct properties of the output.
